# Urinary Peptidomic Profiling in Post-Acute Sequelae of SARS-CoV-2 Infection: A Case-Control Study

**DOI:** 10.1101/2025.10.15.25338065

**Authors:** Dilara Gülmez, Justyna Siwy, Katharina Kurz, Ralph Wendt, Miroslaw Banasik, Björn Peters, Emmanuel Dudoignon, Francois Depret, Mercedes Salgueira, Elena Nowacki, Amelie Kurnikowski, Sebastian Mussnig, Simon Krenn, Samuel Gonos, Judith Löffler-Ragg, Günter Weiss, Harald Mischak, Manfred Hecking, Eva Schernhammer, Joachim Beige, UriCoV working group

## Abstract

**Background:** Post-acute sequelae of severe acute respiratory syndrome coronavirus 2-infection (PASC) is challenging to diagnose and treat, and its molecular pathophysiology remains unclear. Urinary peptidomics can provide valuable information on urine peptides that may enable improved and specified PASC diagnosis.

**Methods:** Using standardized capillary electrophoresis-MS, we examined the urinary peptidomes of 50 patients with PASC 10 months after COVID-19 and 50 controls including healthy individuals (n = 42) and patients with non-COVID-19-associated myalgic encephalomyelitis/chronic fatigue syndrome (n = 8). Based on peptide abundance differences between cases and controls, we developed a diagnostic model using a support vector machine.

**Results:** The abundance of 195 urine peptides among PASC patients significantly differed from that in controls, with a predominant abundance of collagen alpha chains. This molecular signature (PASC195), effectively distinguished PASC cases from controls in the training set [AUC of 0.949 (95% CI 0.900–0.998; p < 0.0001)] and independent validation set [AUC of 0.962 (95% CI 0.897–1.00); p < 0.0001)]. *In silico* assessment suggested exercise, GLP1-RA and MRA as potentially efficacious interventions.

**Conclusions:** We present a novel and non-invasive diagnostic model for PASC. Reflecting its molecular pathophysiology, PASC195 has the potential to advance diagnostics and inform therapeutic interventions.

**Statement of significance of the study:** Despite the recent emergence of omics-derived candidates for post-acute sequelae of SARS-CoV-2 infection (PASC), the pending validation of proposed markers and lack of consensus result in the continuous reliance on symptom-based criteria, being subject to diagnostic uncertainties and potential recall bias. Building upon prior findings of renal involvement in acute COVID-19 pathophysiology and PASC-associated alterations, we hypothesized that the use of urinary peptides for PASC-specific biomarker discovery, unlike conventional specimens that have been utilized thus far, may offer complementary information on putative disease mechanisms. In the present study, 195 significantly expressed peptides were used to form a classifier termed PASC195, which effectively discriminated PASC from non-PASC (p < 0.0001), including healthy individuals and non-COVID-19 associated myalgic encephalomyelitis/chronic fatigue syndrome, in both the derivation (n = 60) and an independent validation set (n = 40). Shift in collagen regulation was associated with PASC, as the majority of PASC195 peptides were derived from collagen alpha chains. Ongoing inflammatory responses, hemostatic imbalances, and endothelial damage were inferred from cross-sectional variations in endogenous peptide excretion.

## 1 Introduction

As of September 21, 2025, severe acute respiratory syndrome coronavirus 2 (SARS-CoV-2) has led to 779 million cumulative COVID-19 cases and 7.1 million associated deaths, with severe health implications for those who have yet to recover fully [1, 2]. Post-acute sequelae of SARS-CoV-2 infection (PASC) or long COVID is a multisystemic chronic condition, characterized by debilitating symptoms that persist or newly arise in the post-acute phase of COVID-19, often presenting with a non-linear, fluctuating clinical course lasting more than three months [3]. Myalgic encephalomyelitis/chronic fatigue syndrome (ME/CFS) can develop after infection with SARS-CoV-2, similar to other infections including those caused by Epstein-Barr virus (EBV) [4, 5]. The diagnosis of both PASC and ME/CFS currently relies on standardized criteria that are strictly limited to clinical, yet inherently subjective, symptom-based assessments [6, 7]. On the other hand, measurable biomarkers have the potential to predict disease onset prior to clinical manifestation, reduce diagnostic ambiguity, and enable both early implementation of therapeutic interventions and longitudinal assessment of therapeutic response. Initially based on demographic and clinical parameters, risk assessment for PASC has progressed from demographic and clinical parameters [8] to five self-reported symptoms in the first week of infection [9] to an interactive recovery prediction tool [10], and more recently, to omics-based biomarkers requiring further validation [11, 12, 13, 14].

SARS-CoV-2 persistence [15], elevated cytokines indicating ongoing inflammation [16], dysregulated complement system [17], and sustained endothelial dysfunction [18, 19, 20] are among the putative pathophysiological mechanisms of PASC, likely engaging in an amplifying feedback loop that drives multi-organ sequelae [21]. Kidneys are among the organs involved due to their relatively strong expression of the cell surface receptor for SARS-CoV-2 (ACE2). A decline in the estimated glomerular filtration rate (eGFR) equivalent to a 3.39% reduction from baseline within the first year following infection and a high prevalence of albuminuria among PASC patients have been described [22]. Proximal tubular injury is characterized by low-molecular-weight proteinuria with aminoaciduria and has also been documented in hospitalized patients with severe COVID-19 [23].

Urinary peptidomic profiling (UPP) is known to reveal disease mechanisms of renal and extra-renal origin, as one-third of urinary proteins are derived from circulating plasma that permeates the glomerular barrier [24]. Capillary-electrophoresis-mass-spectrometry (CE-MS) aided in the discovery of predictive and diagnostic biomarkers through UPP, enabling classifier generation to assess disease mechanisms and progression risk in the subclinical phase of chronic kidney disease [25], diastolic left ventricular dysfunction [26], acute COVID-19 [27], and more recently mortality risk in PASC [28].

Given these observations, we hypothesized that the presence of PASC may be inferred from the urinary excretion of smaller proteins and peptides, particularly endogenous peptides, owing to their intricate implication in pathophysiological mechanisms. Regardless of renal origin, detectable alterations in these peptides make urine a valuable medium for detecting and diagnosing multisystemic diseases like PASC. Here, we implemented a 1:1 matched case-control design, aiming to develop a disease-specific classifier, titled PASC195, through the analysis of urinary peptidomic data from 50 individuals diagnosed with PASC versus their matched controls.

## 2 Materials and Methods

### 2.1 Study design and participants

The cases and controls in our study population originated from several existing cohorts. Figure 1 presents a flowchart of the cohort used to define the potential biomarker, the development, and validation of the classifier. The additional external prospective cohort of patients with previous COVID-19 (UriCoV) is shown in the supplement (Figure E1).

**Figure 1:**
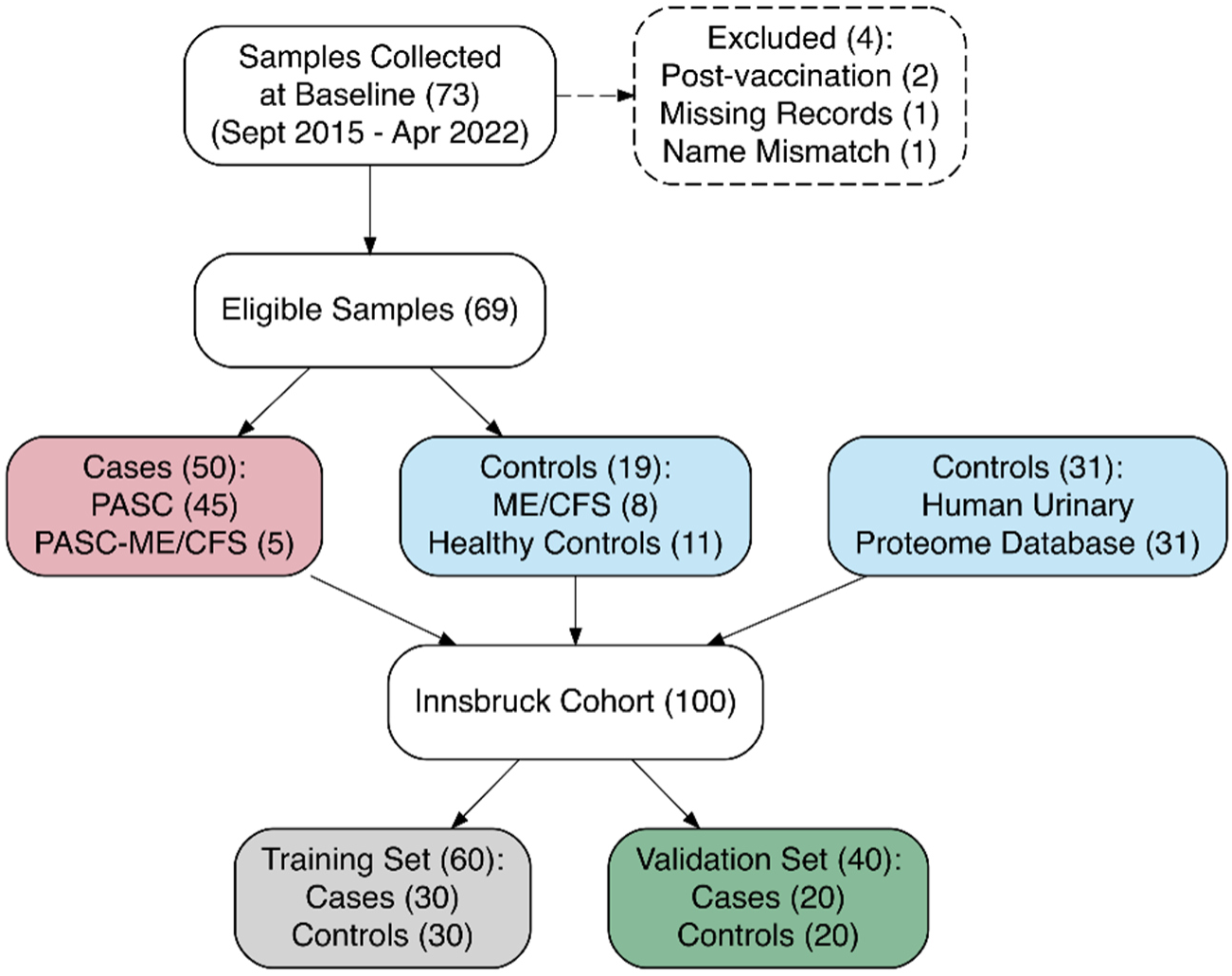
Flowchart for patient recruitment. Legend: Flowchart illustrating the selection process for cases and controls in the Innsbruck cohort. Two patients with symptoms arising post-vaccination without susceptible or confirmed prior to COVID-19 infection, one sample missing both electronic medical records and a completed questionnaire and one case with a name mismatch between the questionnaire and clinical data were excluded from the study. PASC = Post-acute sequelae of SARS-CoV-2 infection. ME/CFS = Myalgic encephalomyelitis/chronic fatigue syndrome.

#### Innsbruck cohort

The participants were sourced from the study, *Establishment of a Blood, Urine, and Stool Sample Archive for Patients with Fatigue After Infection or Recurrent Infections*, conducted at the outpatient clinic for infectious diseases at the Medical University of Innsbruck between September 2015 and April 2022 (ethics vote number 1157/2017). The Innsbruck cohort comprised a total of 100 participants. Self-reported symptoms were quantitatively assessed using the 2003 Canadian Consensus Criteria (CCC) [6] to investigate, whether the patients fulfilled the criteria for ME/CFS. The case group consisted of participants diagnosed with PASC, defined as patients with symptoms persisting for three months after COVID-19 infection. Patients with PASC who also met the 2003 CCC [6] for ME/CFS, without a prior diagnosis of ME/CFS before their COVID-19 infection, were referred to as PASC-ME/CFS. Case urine samples were collected at a median of 292 days (IQR: 177–412) after COVID-19 diagnosis. The control group was selected from healthy individuals and those with non-COVID-19-associated ME/CFS, as verified by electronic medical records (EMRs) that either lacked documentation of previous COVID-19 infection or indicated alternative infectious etiologies. Further controls from the Human Urinary Proteome Database [29] were selected as an extension to 1:1 match the number of cases. The final set of 50 case control pairs was randomly assigned to a training set of pairs (n = 30) or a validation set (n = 20). Demographic and clinical data obtained from the EMRs assisted in matching cases and controls. Age was matched on the calendar year of birth. All participants provided written informed consent and were non-anuric adults (aged ≥18 years).

Detailed descriptions of the *external cohort of previous COVID-19 patients (UriCoV)* used for external evaluation of the biomarker, sample preparation for CE-MS analysis, urinary peptidome analysis, peptide sequencing, and classifier generation are provided in the supplement (Supplementary Methods).

### 2.2 Statistical analysis

A preliminary sample size calculation indicated that 26 case control pairs were required to detect a 40% change in the abundance of a single biomarker, with a type I error of 0.05 and 80% power. Chi-squared and Fisher’s exact tests were used to compare categorical variables. Group differences were assessed using the paired t-test and Wilcoxon rank sum test, respectively, using MedCalc software (version 12.1.0.0; MedCalc Software, Mariakerke, Belgium) and R version 4.4.1 (2024-06-14 ucrt). The description of the statistical analysis performed to define biomarkers is provided in the supplement (Supplementary Methods).

ROC curve analysis assessed the discriminative performance of the classifier within each cohort. The true positive fraction (sensitivity) and false positive fraction (1-specificity) were plotted. The AUC was calculated, and an optimal threshold was established using the Youden index. Exact binomial calculations were used to determine 95% confidence intervals (CI). Symptom duration was measured as the interval between the reported onset of the initial acute COVID-19 infection and the completion of the questionnaire, at which point urine samples were collected or within a short period thereafter.

### 2.3. *In silico* intervention

A recently described pipeline for an *in silico* intervention was applied to the defined specific PASC peptides [30, 31]. The previously described interventions (mineralocorticoid receptor antagonists (MRA), sodium-glucose cotransporter 2 inhibitors (SGLT2i), glucagon-like peptide-1 receptor agonists (GLP-1 RA), angiotensin II receptor blockers (ARBs), olive oil and physical activity) were used both individually and in combination to test which intervention could potentially benefit patients.

### 2.4. Use of artificial intelligence

Artificial intelligence-powered tools leveraging large language models (Research Rabbit, Consensus, ChatGPT OpenAI GPT-4), aided in manuscript preparation for literature research, grammar enhancement, and proofreading. All outputs were reviewed and finalized by the authors.

## 3 Results

### 3.1 Study population

#### Innsbruck cohort

In Innsbruck, 73 urine samples were collected from participants between September 2015 and April 2022 (at the time of diagnosis for ME/CFS or PASC or upon admission for healthy controls), of which 69 were used for UPP (50 cases and 19 controls) (Figure 1). Questionnaire data were available for only 58% of the Innsbruck cohort (94% of cases and 42.1% of controls), likely because of secondary extension of the control group and non-participation among asymptomatic controls. The potential non-participation bias was mitigated by incorporating detailed clinical diagnostic information obtained from electronic medical records for all participants. This ensured that group allocation was based not only on self-reported symptoms but also on clinician-confirmed diagnoses, thereby supporting the robustness of case–control classification. Further datasets of sex- and age-matched individuals were extracted from the Human Urinary Proteome Database. The sample characteristics of the Innsbruck cohort are presented in Table 1.

**Table 1:**
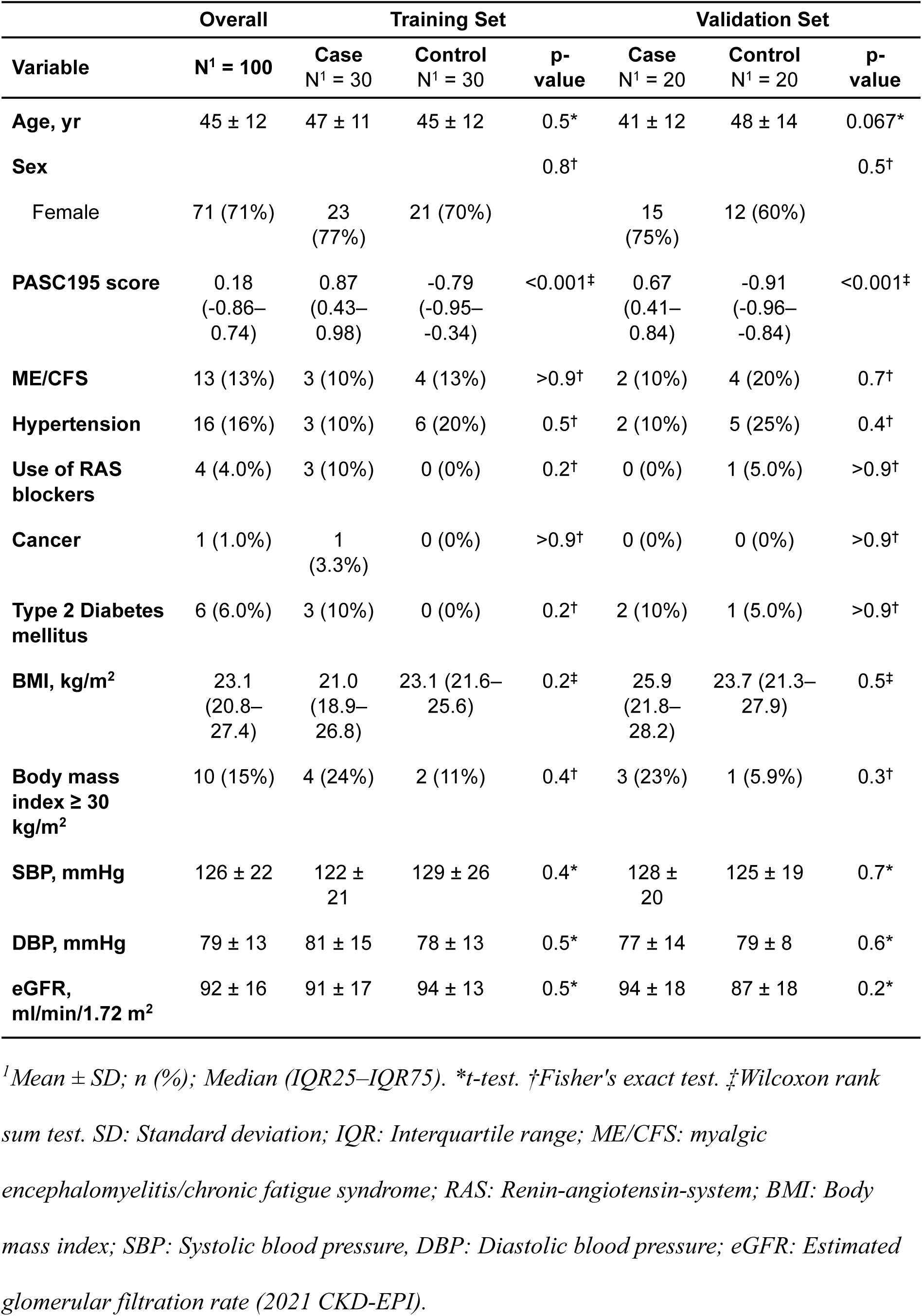
Characteristics – Innsbruck Cohort.

| Variable | Overall | Training Set |  |  | Validation Set |  |  |
| --- | --- | --- | --- | --- | --- | --- | --- |
|  | N<sup>1</sup> = 100 | Case<br>N<sup>1</sup> = 30 | Control<br>N<sup>1</sup> = 30 | p-<br>value | Case<br>N<sup>1</sup> = 20 | Control<br>N<sup>1</sup> = 20 | p-<br>value |
| **Age, yr** | 45 ± 12 | 47 ± 11 | 45 ± 12 | 0.5* | 41 ± 12 | 48 ± 14 | 0.067* |
| **Sex** |  |  |  | 0.8<sup>†</sup> |  |  | 0.5<sup>†</sup> |
| Female | 71 (71%) | 23<br>(77%) | 21 (70%) |  | 15<br>(75%) | 12 (60%) |  |
| **PASC195 score** | 0.18<br>(-0.86–<br>0.74) | 0.87<br>(0.43–<br>0.98) | -0.79<br>(-0.95–<br>-0.34) | <0.001<sup>‡</sup> | 0.67<br>(0.41–<br>0.84) | -0.91<br>(-0.96–<br>-0.84) | <0.001<sup>‡</sup> |
| **ME/CFS** | 13 (13%) | 3 (10%) | 4 (13%) | >0.9<sup>†</sup> | 2 (10%) | 4 (20%) | 0.7<sup>†</sup> |
| **Hypertension** | 16 (16%) | 3 (10%) | 6 (20%) | 0.5<sup>†</sup> | 2 (10%) | 5 (25%) | 0.4<sup>†</sup> |
| **Use of RAS**<br>**blockers** | 4 (4.0%) | 3 (10%) | 0 (0%) | 0.2<sup>†</sup> | 0 (0%) | 1 (5.0%) | >0.9<sup>†</sup> |
| **Cancer** | 1 (1.0%) | 1<br>(3.3%) | 0 (0%) | >0.9<sup>†</sup> | 0 (0%) | 0 (0%) | >0.9<sup>†</sup> |
| **Type 2 Diabetes**<br>**mellitus** | 6 (6.0%) | 3 (10%) | 0 (0%) | 0.2<sup>†</sup> | 2 (10%) | 1 (5.0%) | >0.9<sup>†</sup> |
| **BMI, kg/m<sup>2</sup>** | 23.1<br>(20.8–<br>27.4) | 21.0<br>(18.9–<br>26.8) | 23.1 (21.6–<br>25.6) | 0.2<sup>‡</sup> | 25.9<br>(21.8–<br>28.2) | 23.7 (21.3–<br>27.9) | 0.5<sup>‡</sup> |
| **Body mass**<br>**index ≥ 30**<br>**kg/m<sup>2</sup>** | 10 (15%) | 4 (24%) | 2 (11%) | 0.4<sup>†</sup> | 3 (23%) | 1 (5.9%) | 0.3<sup>†</sup> |
| **SBP, mmHg** | 126 ± 22 | 122 ±<br>21 | 129 ± 26 | 0.4* | 128 ±<br>20 | 125 ± 19 | 0.7* |
| **DBP, mmHg** | 79 ± 13 | 81 ± 15 | 78 ± 13 | 0.5* | 77 ± 14 | 79 ± 8 | 0.6* |
| **eGFR,**<br>**ml/min/1.72 m<sup>2</sup>** | 92 ± 16 | 91 ± 17 | 94 ± 13 | 0.5* | 94 ± 18 | 87 ± 18 | 0.2* |

<sup>1</sup>Mean ± SD; n (%); Median (IQR25–IQR75). \*t-test. <sup>†</sup>Fisher's exact test. <sup>‡</sup>Wilcoxon rank sum test. SD: Standard deviation; IQR: Interquartile range; ME/CFS: myalgic encephalomyelitis/chronic fatigue syndrome; RAS: Renin-angiotensin-system; BMI: Body mass index; SBP: Systolic blood pressure, DBP: Diastolic blood pressure; eGFR: Estimated glomerular filtration rate (2021 CKD-EPI).

The mean age of the participants was 45.3 years, and 71% were female. Comorbidities included type 2 diabetes mellitus (n = 6), overweight (n = 10), cancer (n = 1), and hypertension (n = 16) with four patients being on Renin-Angiotensin System (RAS) blockade therapy. One patient had both diabetes and hypertension. Of the eight non-COVID-19 ME/CFS patients, two had documentation of chronic or previous EBV infection, one patient had both EBV and Toxocara infection, one had documented Ehrlichiosis, and four were classified as having an unclear etiology. At baseline, 13% of the Innsbruck cohort fulfilled the CCC for ME/CFS [6], compared to 10% of those with PASC.

### 3.2 Classifier generation and discriminatory power of PASC195

Training data from the Innsbruck cohort, including 30 cases (27 PASC, 3 PASC-ME/CFS) and 30 controls (26 healthy controls, 4 non-COVID-19 ME/CFS) were used to define the PASC-associated urinary peptides. Of the 2,034 peptides detected in the urine peptidome analysis, 375 differed significantly in abundance (Figure 2, Panel A). Amino acid sequences could be obtained for 243 peptides, which were further subjected to a take-one-out process. This refinement resulted in a final pool of 195 peptides used to generate a classifier, PASC195, consisting of 187 upregulated and eight downregulated peptides. Of these, 172 (88.2%) were aligned to collagen alpha, with 132 (76.7%) linked to collagen alpha-1 and 29 (16.9%) to collagen alpha-2 chains. Other peptides included fragments of alpha-1-antitrypsin (A1AT), antithrombin-III (AT III), apolipoprotein A-II (APOA2), apolipoprotein A-IV (APOA4), basement membrane-specific heparan sulfate proteoglycan core protein (PGBM), beta-2-microglobulin (B2M), calreticulin (CALR), CD99 antigen, fibrinogen alpha (FIBA) and beta (FIBB), serotransferrin (TF), protein S100-A9, and mannan-binding lectin serine protease 2 (MASP2), among others. Among the upregulated peptide sequences, the majority were associated with collagen alpha-1(I) (CO1A1) (n = 50), collagen alpha-1(III) (CO3A1) (n = 29) and collagen alpha-2(I) (CO1A2) (n = 13). Downregulated peptides were fragments of collagen alpha-2(IV) (CO4A2) (n = 2), collagen alpha-1(XIV) (COEA1) (n = 1), clathrin (CLCB) (n = 1), glycogen synthase kinase-3 alpha (GSK3A) (n = 1), and Golgin subfamily A member 6-like protein 2 (GOLGA6L2) (n = 1), among others.

**Figure 2:**
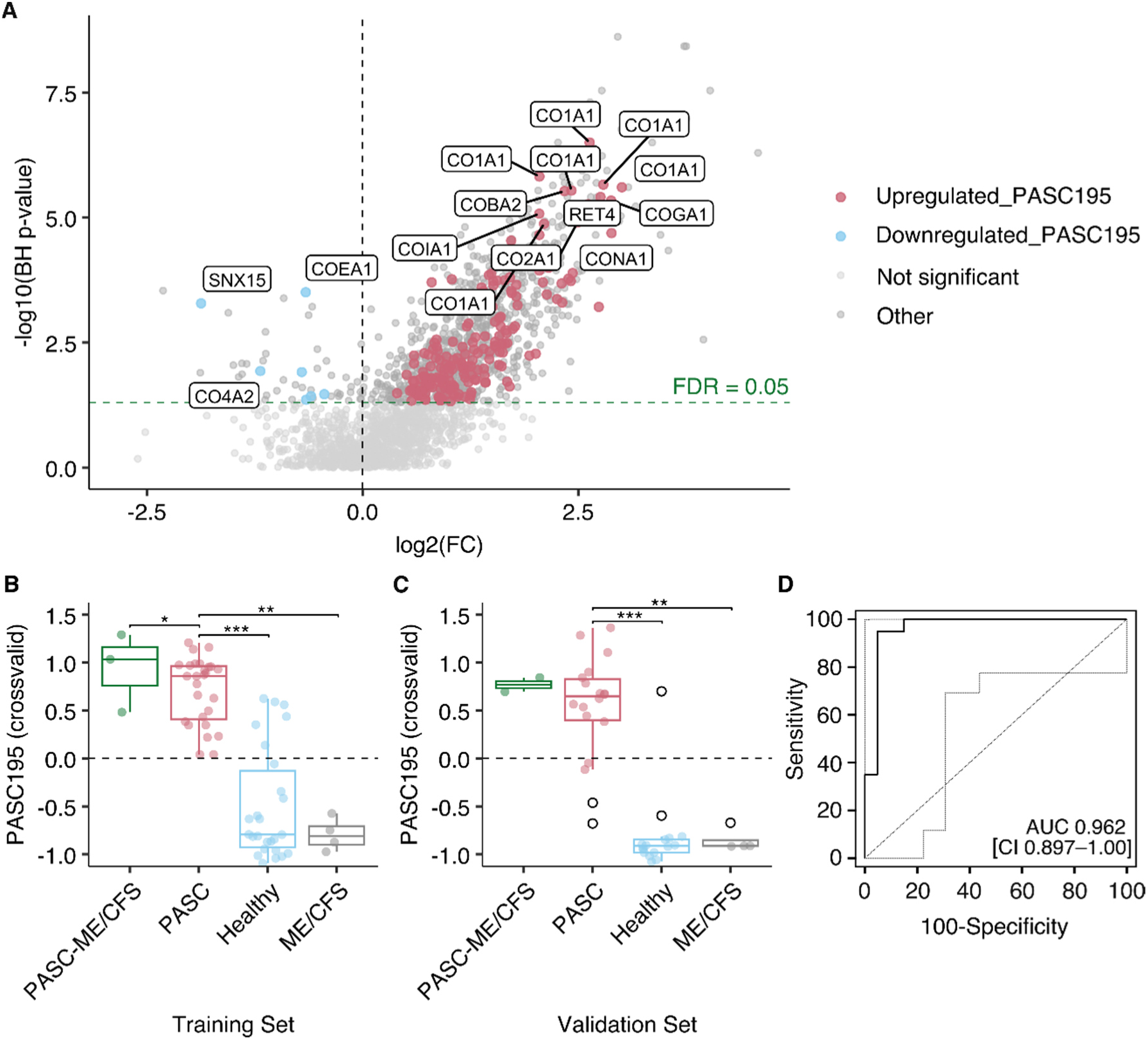
Classifier generation and performance of PASC195. Legend: Panel A: Volcano plot displaying log2 fold change (FC) in peptide abundance. FC was calculated as the mean peptide abundance within cases divided by the mean in the control group. The horizontal line indicates the significance threshold (FDR = 0.05). Red dots represent significantly upregulated peptides in PASC195 and blue dots represent significantly downregulated peptides. Light gray indicates non-significant peptides and gray other peptides. Panel B: PASC195 classification scores of the training set are shown as box-whisker plots for PASC-ME/CFS (n = 3), PASC (n = 27), healthy controls (n = 26), and non-COVID-19 ME/CFS (n = 4). Pairwise comparisons between groups were conducted using the Wilcoxon rank sum test. Asterisks (*) indicate p < 0.05, (**) indicate p < 0.01, and (***) indicate p < 0.001 (BH-corrected). Panel C: PASC195 classification scores of the validation set are shown as box-whisker plots for PASC-ME/CFS (n = 2), PASC (n = 18), healthy controls (n = 16) and ME/CFS within the control group (n = 4). Black circles indicate outliers. Panel D: Receiver Operating Characteristic (ROC) curve visualizing the performance of PASC195 in distinguishing between cases (n = 20) and controls (n = 20) within the validation set. The curve plots the sensitivity against 100 -specificity, achieving an AUC of 0.962 (95% CI 0.897–1.00; p < 0.0001). The bold black line represents the ROC curve. The gray dotted lines represent 95% confidence intervals. PASC: Post-acute sequelae of SARS-CoV-2 infection. ME/CFS: Myalgic encephalomyelitis/chronic fatigue syndrome.

The median cross-validated score in the case group was 0.74 (IQR: 0.44 to 0.96) and in the controls, -0.86 (IQR: -0.94 to -0.61), respectively. PASC195 achieved an AUC of 0.949 (95% CI 0.900–0.998; p < 0.0001) when estimated using complete leave-one-out cross-validation on the training set (Figure 2, Panel B). In the independent validation set of 40 individuals (20 cases and 20 controls), the classifier effectively distinguished patients with PASC from controls, yielding an AUC of 0.962 (95% CI 0.897–1.00; p < 0.0001) (Figure 2, Panels C-D). The sensitivity and specificity of 95% were at a cut-off of -0.596 (positive and negative predictive value of 95%), supporting the model’s robustness and predictive accuracy in an independent dataset. The peptidome profile of PASC195 is shown in Figure 3 and the Supplemental Results (E3).

**Figure 3:**
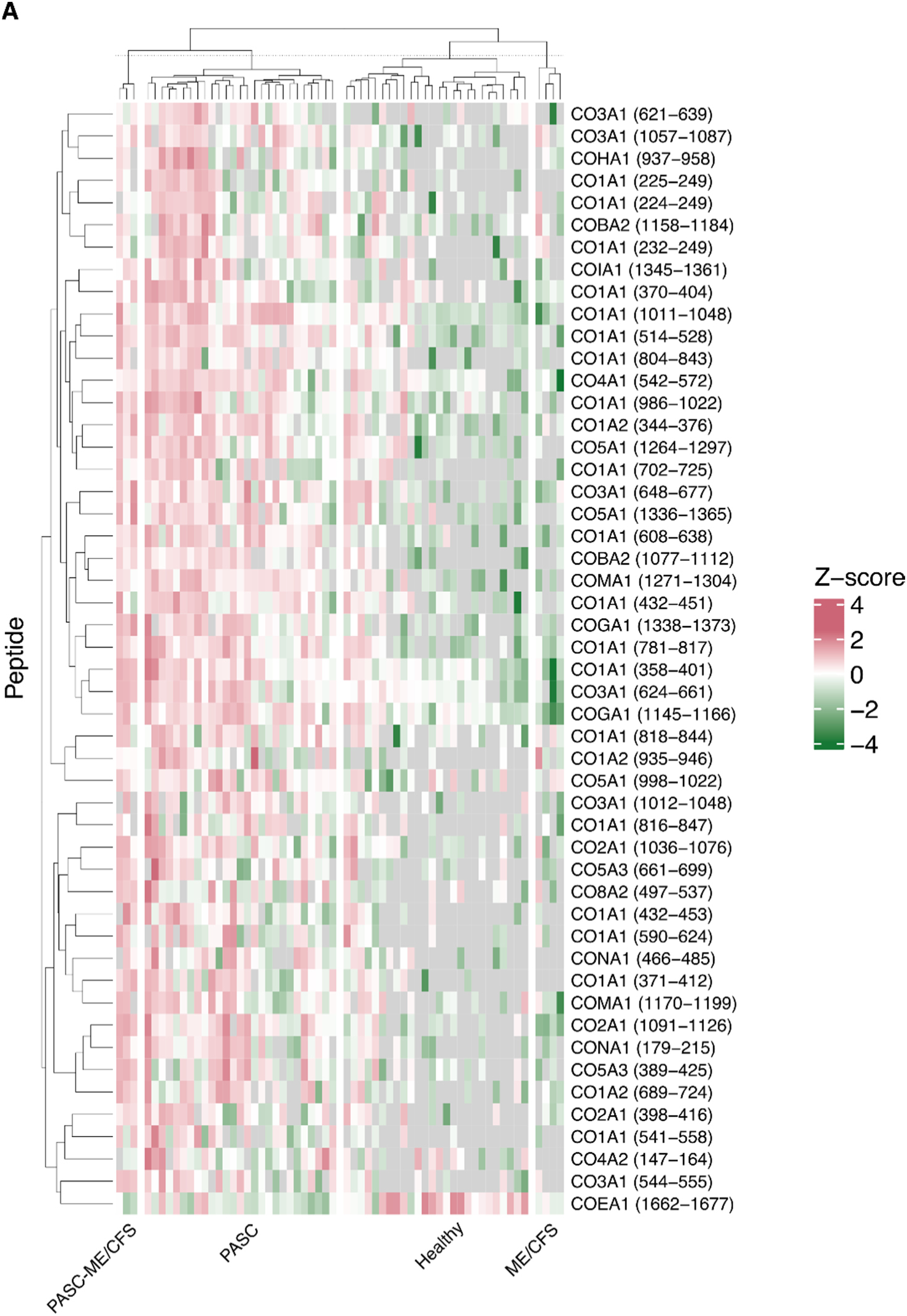
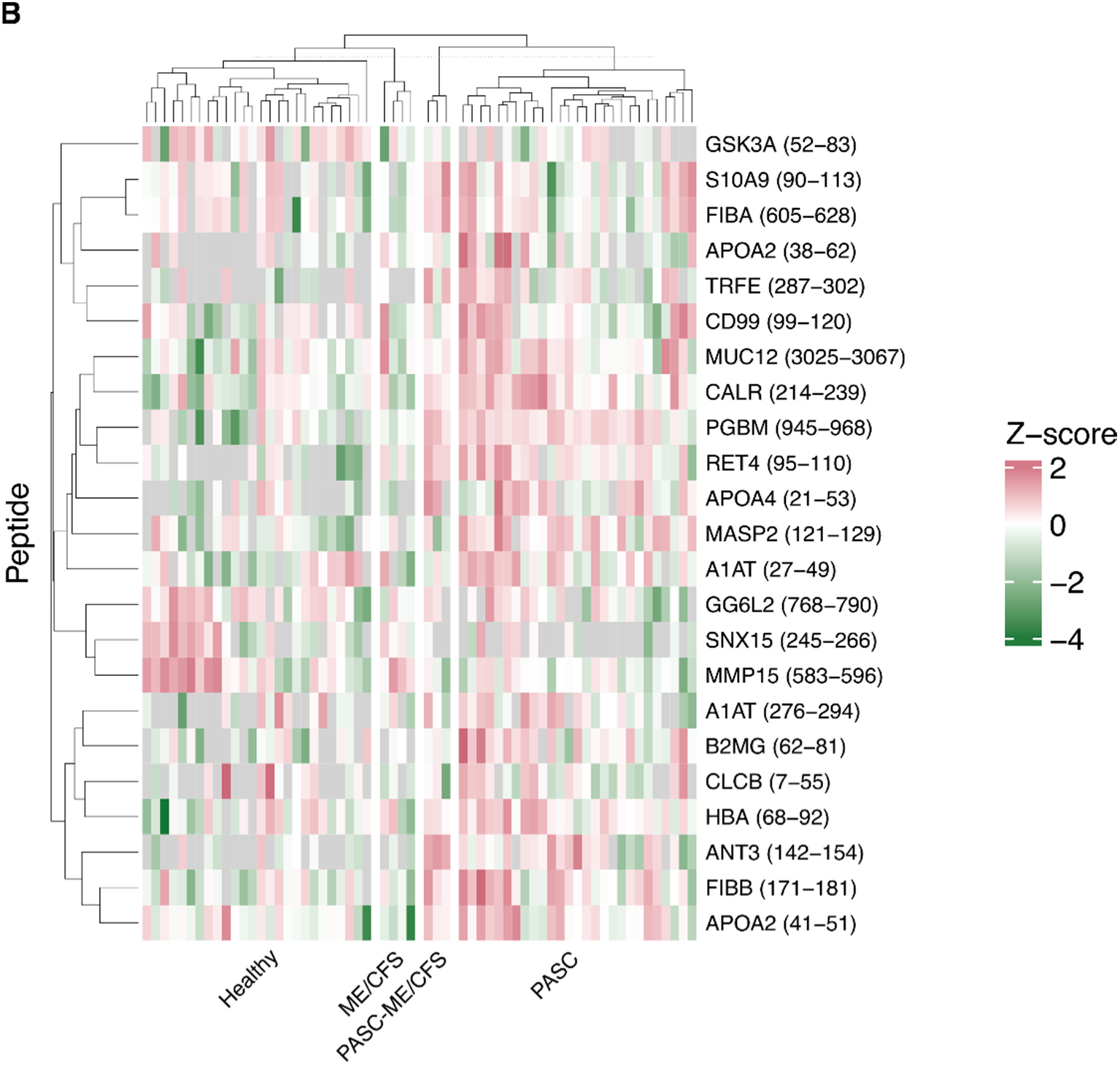
Urinary peptidomics profiling (UPP) of the training set by subgroup (PASC195) Legend: UniProt symbols corresponding to each peptide are displayed along the y-axis. Log2-transformed peptide signal intensities were Z-normalized and hierarchically clustered within four subgroups: PASC-ME/CFS, PASC, healthy controls, and non-COVID-19 ME/CFS. The start and end positions of the amino acids are indicated in parentheses. Panel A displays the top 50 significant collagenous peptides, and panel B illustrates all the non-collagenous peptides of the PASC195 classifier. See supplementary material for the complete peptidome profile. Green, pink, and gray indicate higher signal intensities than mean peptide abundance, lower signal intensities, and missing values, respectively. PASC: Post-acute sequelae of SARS-CoV-2 infection. ME/CFS: Myalgic encephalomyelitis/chronic fatigue syndrome.

### 3.3 *In silico* intervention

As no specific therapeutic approach has yet demonstrated consistent efficacy in PASC, we employed our recently described algorithm to simulate *in silico* responses to six interventions (MRA, SGLT2i, GLP-1 RA, ARB, olive oil and physical activity) [30, 31]. The algorithm, applied to the PASC patient data, estimated heterogeneous individual responses (Supplemental Table E2). The average predicted improvement in the PASC195 scores was 0.131 ± 0.044 when the most suitable intervention was assigned to each patient. Physical activity, MRA and GLP-1 RA, in particular, were predicted to be beneficial in most cases.

### 3.4 Clinical symptoms in case and control subgroups and their association with PASC195 score

Self-reported symptoms at baseline [median duration: 300 days (IQR: 169–384)] were assessed using CCC [6], which categorized the diagnostic criteria for ME/CFS into seven domains and 38 symptoms. Concentration impairment and short-term memory consolidation were the most frequently reported symptoms (74.1%), as were by cases [80.9%, a median duration of 303 days (IQR: 186–426)]. Tender lymph nodes were the least reported symptom (13.8%), with a median duration of 325 days (IQR: 220–334). Detailed response distributions for each symptom are presented in Figure 4 and the supplement (Supplementary Table E3).

**Figure 4:**
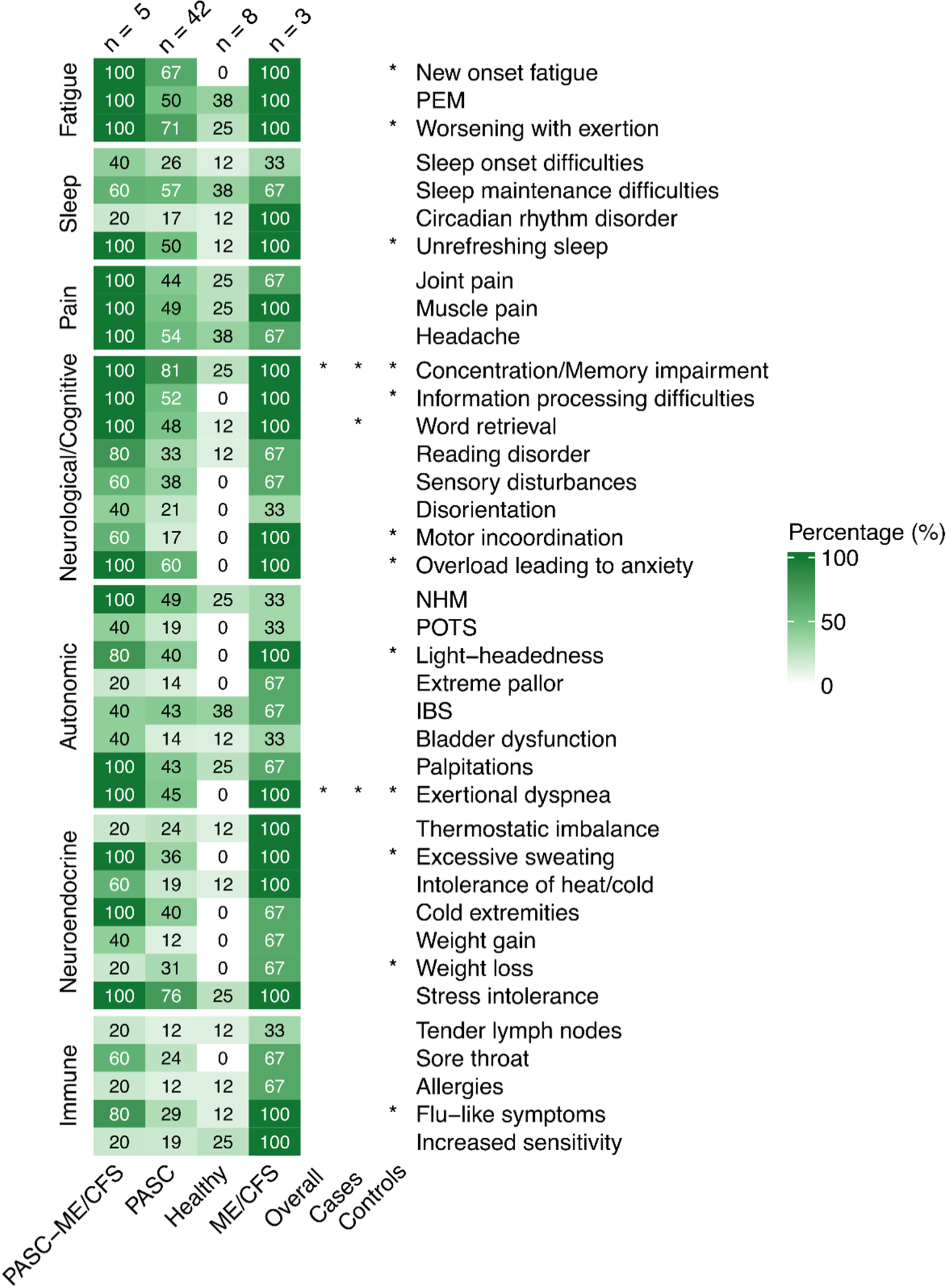
Symptom profile by diagnosis and associations with PASC195 scores. Legend: Heatmap illustrating response distributions in percentages (%) for four subgroups (PASC-ME/CFS, PASC, healthy controls, and non-COVID-19 ME/CFS) by manifestation. The color intensity increases by percentages. The sample sizes of questionnaire respondents are shown in parentheses. Significant differences in the classification scores between symptomatic and asymptomatic patients are marked with asterisks (*) in the overall cohort, cases, and controls (Wilcoxon rank sum test). P-values were not corrected for multiple testing. (*) p < 0.05, (**) p < 0.01, and (***) p < 0.001. ME/CFS: Myalgic encephalomyelitis/chronic fatigue syndrome; PASC: Post-acute sequelae of SARS-CoV-2 infection. PEM: Post-Exertional Malaise. NHM: Delayed postural hypertension or neurally mediated hypotension. POTS: Postural orthostatic tachycardia syndrome. IBS: Irritable bowel syndrome.

Significantly higher PASC195 scores were found in the presence of concentration impairment and short-term memory consolidation (r = 0.27, p = 0.044), and exertional dyspnea (r = 0.26, p = 0.048) among all questionnaire respondents, as shown in Figure 4. In a subgroup analysis of only cases, significant differences were found in word retrieval (r = 0.31, p = 0.036), concentration impairment and short-term memory consolidation (r = 0.32, p = 0.031), and exertional dyspnea (r = 0.31, p = 0.032), with each symptom associated with higher scores compared to cases without the respective symptom. However, due to the exploratory nature of this analysis, these calculations were not corrected for multiple testing.

#### UriCoV cohort

The classifier’s performance was evaluated in the multicentric UriCoV prospective cohort. Sensitivity analysis yielded no statistically significant difference (p = 0.1492) between cases (n = 34) and controls (n = 40). Detailed findings are included in the supplement (Supplemental Results).

## 4 Discussion

In the absence of consensus-based diagnostic models derived from an understanding of disease mechanisms, we aimed to identify urinary peptides hinting at PASC pathophysiology using a data-driven approach. While direct comparisons between our urinary peptidome data and previously reported plasma or serum peptidomes are not feasible due to differences in sample types, methodologies, and data processing, we observed analogous patterns. We found molecular patterns of collagen dysregulation, prothrombotic events, and potential signs of endothelial dysfunction reflected in the urine peptides to be associated with PASC, consistent with similar findings reported in previous omics-based PASC studies using serum or plasma as derivatives [32, 17, 33, 34, 35, 12]. Second, we developed a novel classifier, PASC195, comprising 195 modulated peptides that significantly differed from the UPP of individuals without PASC, including other post-acute infectious syndromes progressing to ME/CFS. Third, an exploratory analysis of classification scores and 38 symptoms linked PASC195 to neurocognitive (word retrieval deficits, concentration impairment and short-term memory consolidation), as well as autonomic manifestations (exertional dyspnea).

UPP captures peptides originating from plasma as well as renal or urinary tract; however, assigning a specific origin to individual peptides is technically complex. The dysregulated peptides observed in PASC likely arise from a combination of systemic and renal processes, including inflammation, endothelial dysfunction, and potentially altered proteolysis. While our data did not permit quantification of proteolytic activity per se (increased proteolysis), the associations between PASC195 scores and neurocognitive as well as autonomic symptoms suggest that the urinary peptidome reflects multisystem involvement characteristic of PASC. Future studies integrating urine, plasma, and tissue-level proteomics will be essential to determine the origin and mechanistic relevance of these peptides.

A shift in collagen homeostasis is a representative feature of PASC195, as the majority of dysregulated peptide sequences stem from CO1A1, like the COV50 classifier, a urinary proteomic biomarker comprising 50 peptides. COV50 has been associated with predicting adverse outcomes during the acute phase and a predisposition to death even in the absence of COVID-19 infection [36]. The CO1A1 downregulation in COV50 has been attributed to attenuated collagen degradation within the extracellular matrix, likely leading to increased fibrosis [36]. In contrast to COV50, CO1A1 fragments were upregulated in PASC195, with no overlapping in peptide sequences between the two biomarkers. Another mutual component of COV50 and PASC195 are the degradation products of A1AT, which were found to be upregulated in COVID-19 [27], and here in PASC, respectively. A1AT is an inhibitor of serine proteases, like TMPRSS-2, which assists SARS-CoV-2 in binding to the ACE2 receptor via spike protein activation and also acts as an acute-phase protein with anti-inflammatory and vascular protective effects [37]. Its higher abundance could imply an amplification loop of ongoing inflammation or endothelial damage in PASC.

Confirming earlier findings from a study on the plasma proteome in patients with PASC [34, 35, 12], we detected upregulation of fibrinogen - FIBA and FIBB - in the urine peptidome. Notably, the same study demonstrated decreased FIBA and FIBB levels in pellet fractions. One possibility is that this reduction results from abnormalities in proteolytic activity or accelerated degradation, which may manifest as a consequent enrichment of fibrinogen subunits in urine. A recent study on the serum proteome identified elevated levels of FIBB alongside other factors of the coagulation cascade (coagulation factor XI, protein C, and heparin cofactor II), as well as decreased AT III (linked by the authors to increased cleavage activity, potentially explaining our findings regarding its abundance) at 6 months post-infection [17]. Because fibrin is involved not only in platelet aggregation but also in preventing IFN-γ induced hemorrhage and triggering immune responses [38], these findings should not be confined to a hypercoagulable state alone. Disruptions in iron homeostasis and inflammatory anemia were described as early indicators of PASC in a longitudinal cohort study [39]. Serum iron and transferrin saturation persisted at significantly lower levels for up to nine months after severe/critical COVID-19, consistent with our findings regarding the high prevalence of impaired cognitive function, fatigue, and upregulation of serum transferrin observed.

Siwy et al. reported CD99 antigen depletion specific to severe acute COVID-19 in a study on urine peptidome [40]. Given its role in leukocyte extravasation and T-cell integration [38], a compensatory immune response likely accounts for its upregulation in PASC. Calprotectin (S100A8/A9) has been shown to correlate with disease severity and increased mortality risk in acute COVID-19 [32, 41, 12]. In line with immune dysregulation, we found a higher abundance of peptides derived from calprotectin subunits (S100A9). Another remarkable finding was the overexpression of PGBM, a component of basal membranes including the glomerular basement membrane, possibly linked to increased peptide leakage or systemic alterations in endothelial barrier function.

Following our recently published approach for *in silico* intervention in CKD [31], we applied the algorithm to the PASC patient data described here. This exploratory analysis, motivated in part by the lack of established therapies for PASC, identified exercise, MRA, and GLP-1 Receptor agonists as potentially beneficial for the majority of patients. While this approach does not constitute evidence of efficacy (and some interventions, especially exercise, may not be feasible for all), it outlines a potential path forward for targeted interventions.

Despite the fluctuating course of PASC, symptoms have often been observed to resolve or improve over the subsequent years [42]. Urine sampling in the UriCoV cohort occurred approximately three years after the initial SARS-CoV-2 infection, compared to ten months post-infection in the Innsbruck cohort. In UriCoV, the presence of ongoing or prior symptoms at the time of sampling was assessed through a structured self-report questionnaire, primarily reflecting participants’ perceived symptom burden, as clinical evaluation was not part of the study protocol, whereas in Innsbruck, case status was established through clinician-confirmed diagnosis. The high proportion of reported cases in UriCoV indicates that differences in questionnaire design and reliance on self-reported data may have contributed to diagnostic variability, potentially affecting consistency in case classification. Unlike the Innsbruck participants, UriCoV subjects were not routinely recruited from an infectious disease outpatient clinic, which may have introduced selection bias related to symptom perception and reporting. Taken together, these factors, recognized only during the study, suggest that at the time of sampling, the clinical presentation of participants classified as PASC in UriCoV likely differed in severity or stage from those examined ten months post-infection in Innsbruck. Cohort-specific differences in case and control definitions, symptom burden, and participant selection likely contributed to variability in urinary peptidomic profiles and precluded validation of the PASC195 classifier in this independent cohort, thereby limiting its generalizability. These inconsistencies underscore the need for harmonized diagnostic criteria and standardized recruitment and phenotyping procedures in future studies to enable cross-cohort comparability and support validation of urinary biomarkers in broader PASC populations.

Moreover, including individuals with other post-acute infectious syndromes as a separate comparator group, as well as conducting sub-cohort analyses by sex, age, or other clinical parameters, would have aided in identifying potential subgroup-specific associations and in delineating PASC-specific molecular signatures. However, the limited sample size did not permit such analyses, as the small number of participants in each subgroup would have substantially reduced statistical power and reliability. We therefore acknowledge this as a limitation of the current study. Future investigations with larger and more heterogeneous populations will be essential to explore potential sex- or age-specific differences in urinary peptidomic alterations associated with PASC. Future studies with longitudinal designs, enrolling diverse post-infectious cohorts and integrating urine and plasma peptidomics, will be key to confirming these findings and elucidating the mechanisms underlying PASC.

In summary, we present a PASC-specific urinary peptidome profile that revealed cross-sectional variations in collagen homeostasis, inflammatory responses, and hemostatic processes that are distinct from non-COVID-19 ME/CFS and healthy controls, informing differences in disease pathogenesis. Our findings not only hold clinical and predictive relevance by enabling PASC diagnosis at a molecular level but also provide an objective biological correlate that may complement patient-reported outcomes. This has established the key prerequisites for more precise detection and, through *in silico* intervention, a potential pathway toward the treatment of PASC.

## 5 Associated Data

Patient data cannot be made publicly available due to GDPR restrictions. The datasets used and/or analyzed during the current study are available from the corresponding author on reasonable request. Proposals will be reviewed and approved by the authors with scientific merit and feasibility as the criteria. After approval of a proposal, data can be shared via a secure online platform after signing a data access and confidentiality agreement. Data will be made available for a maximum of 5 years after a data sharing agreement has been signed.

## Data Availability

All data produced in the present study are available upon reasonable request to the authors

## Acknowledgements

UriCoV Working Group: Justyna Siwy, Mosaiques Diagnostics GmbH, Hannover, Germany; Ralph Wendt, Department of Nephrology, St. Georg Hospital, Leipzig, Germany; Joachim Beige, Division of Nephrology, St. Georg Hospital, Leipzig, Department of Internal Medicine II, Martin-Luther-University Halle-Wittenberg, Halle, Germany, and Kuratorium for Dialysis and Transplantation (KfH) Leipzig, Leipzig, Germany; Miroslaw Banasik, Department of Nephrology , Transplantation Medicine and Internal Diseases Wrocław Medical University, Wroclaw, Poland; Björn Peters, Department of Molecular and Clinical Medicine, Institute of Medicine, the Sahlgrenska Academy at University of Gothenburg, Gothenburg, Sweden, and Department of Nephrology, Skaraborg Hospital, Skövde, Sweden; Emmanuel Dudoignon, Hospital Saint Louis-Lariboisière, Paris, France; Dilara Gülmez, Lenka Grula, Amelie Kurnikowski, and Manfred Hecking, all from the Department of Epidemiology, Medical University of Vienna, Vienna, Austria;,; Andrzej Konieczny, Magdalena Krajewska, Justyna Zachciał, Dorota Bartoszek, Patryk Wawrzonkowski, and Krzysztof Wiśnicki from the Department of Nephrology, Transplantation Medicine and Internal Diseases , Marta Kepinska from Department and Institute of Pharmaceutical Biochemistry, Wrocław Medical University, Wroclaw, Poland; Emelie Sarenmalm, Region Västra Götaland, Skaraborg Hospital, Department of Infectious Diseases, Skövde, Sweden; Åsa Nilsson, Region Västra Götaland, Skaraborg Hospital, Research, Education, Development, and Innovation Department, Skövde, Sweden; Goce Spasovski, University Sts. Cyril and Methodius, Skopje, Republic of North Macedonia; Mercedes Salgueira Lazo, Virgen Macarena Hospital and University of Seville, Seville; Maria Isabel García Sánchez, Biobank Node at Virgen Macarena Hospital, Seville, integrated in the Spanish National Biobanks Network (PT23/00134); Marek W Rajzer from the First Department of Cardiology, Interventional Electrocardiology and Arterial Hypertension, Jagiellonian University Medical College, Kraków, Poland; Beata Czerwieńska, from the Department of Nephrology, Endocrinology, and Metabolic Diseases, Medical University of Silesia, Katowice, Poland; Magdalena Dzitkowska—Zabielska from the Faculty of Physical Education, Gdańsk University of Physical Education and Sport and Centre of Translational Medicine, Medical University of Gdańsk, Gdańsk, Poland; Łukasz Fuławka from Molecular Pathology Centre Cellgen, Wrocław, Poland; Elena Nowacki, University of Patients–Sorbonne University, Paris, France; Catherine Tourette-Turgis, University of Patients, research chair “Compétences & vulnérabilités” Sorbonne University, France; Morgane Michel, Université Paris Cité, ECEVE, UMR 1123, Inserm, Paris, France; Assistance Publique-Hôpitaux de Paris, Hôpital Robert Debré, Unité d’épidémiologie clinique, Paris, France.

## List of abbreviations

A1AT: Alpha-1-antitrypsin
ARB: Angiotensin II receptor blockers
APOA2: Apolipoprotein A-II
APOA4: Apolipoprotein A-IV
AT: III Antithrombin-III
AUC: Area under the curve
B2M: Beta-2-microglobulin
CALR: Calreticulin
CCC: Canadian Consensus Criteria
CE-MS: Capillary-electrophoresis-mass-spectrometry
CI: Confidence intervals
CLCB: Clathrin
CO1A1: Collagen alpha-1(I)
CO1A2: Collagen alpha-2(I)
CO3A1: Collagen alpha-1(III)
CO4A2: Collagen alpha-2(IV)
COEA1: Collagen alpha-1(XIV)
COVID-19: Coronavirus disease 2019
EBV: Epstein-Barr virus
eGFR: Estimated glomerular filtration rate
EMRs: Electronic medical records
FIBA: Fibrinogen alpha
FIBB: Fibrinogen beta
GLP-1: R Glucagon-like peptide-1 receptor agonists
GOLGA6L2: Golgin subfamily A member 6-like protein 2
GSK3A: Glycogen synthase kinase-3 alpha
IBS: Irritable bowel syndrome
IQR: Interquartile range
MASP2: Mannan-binding lectin serine protease 2
ME/CFS: Myalgic encephalomyelitis/chronic fatigue syndrome
MRA: Mineralocorticoid receptor antagonists
NHM: Delayed postural hypertension or neurally mediated hypotension
PASC: Post-acute sequelae of SARS-CoV-2 infection
PEM: Post-exertional malaise
PGBM: Basement membrane-specific heparan sulfate proteoglycan core protein
POTS: Postural orthostatic tachycardia syndrome
ROC: Receiver operating characteristic
S100A8/A9: Calprotectin
SARS-CoV-2: Severe acute respiratory syndrome coronavirus 2
SGLT2i: Sodium-glucose cotransporter 2 inhibitors
TF: Serotransferrin
UPP: Urinary peptidomic profiling

## Data Supplement

Supplemental Methods: PASC-Specific Biomarker Definition

Supplemental Results: UriCoV Validation Set

Supplementary Table E1: Classification and Urine Sample Collection (UriCoV)

Supplementary Table E2: *In silico* intervention results.

Supplementary Table E3: Response distribution and symptom duration among questionnaire respondents

Supplementary Table E4: Statistical Comparison of Subgroups

Supplementary Table E5: Associations with Peptidomics-Derived Classification Scores

Figure E1: Flowchart for the UriCoV cohort

Figure E2: Sensitivity analysis in the validation set of UriCoV

Figure E3: Peptidome profile of all collagenous peptides in the PASC195 classifier

## Declarations

### Ethics approval and consent to participate

Medical University of Innsbruck - Ethics vote number 1157/2017; German-Saxonian Board of Physicians (Dresden, Germany; number EK-BR-88/20.1). Informed consent was obtained from all subjects involved in this study.

## Authors’ contributions

Conceptualization: J.S, H.M, J.B, M.H, R.W, B.P, M.B; Methodology: J.S, H.M; Formal Analysis: J.S, H.M, D.G; Investigation: D.G, J.S, H.M, J.B, M.H, R.W, B.P, M.B, A.M, K.K, E.N; Resources: J.S, H.M, J.B, M.H, R.W, B.P, S.M, A.M; Data collection: D.G, K.K, J.S, S.G, S.E; Writing – original draft: D.G; Writing – review and editing: J.S, H.M, M.H, A.K, S.K, S.M, J.B, B.P, E.D, M.B, K.K, G.W, J.R; Visualization: D.G; J.S; Supervision: J.S, H.M, KK, M.H, J.B; Project administration: J.S; Funding acquisition: H.M, M.H

