## Supplementary material for "Urinary Peptidomic Profiling in Post-Acute Sequelae of SARS-CoV-2 Infection: A Case-Control Study"

### Contents

1 Supplemental Methods ... 3

1.1 Study design - UriCoV ... 3

1.2 PASC-specific biomarker definition ... 4

1.2.1 Sample preparation ... 4

1.2.2 Urinary peptidome analysis ... 5

1.2.3 Sequencing of peptides ... 5

1.2.4 Statistical analysis and biomarker classifier generation ... 6

2 Supplemental Results ... 7

2.1 UriCoV cohort ... 7

2.2 Application of PASC195 to the UriCoV cohort ... 7

3 Supplemental References ... 8

4 Supplemental Tables & Figures ... 10

### 1 Supplemental Methods

#### 1.1 Study design - UriCoV

*External cohort of previous COVID-19 patients.* Further prospective urine samples were collected in the frame of the *URInary peptidomic patterns of Long-COVID syndrome (UriCoV)*, which builds on our previous European multicenter cohort study, *Prospective validation of a proteomic urine test for early and accurate prognosis of critical course complications in patients with SARS-CoV-2 infection* or Crit-COV-U [1]. Ethics approval for UriCoV was obtained via an amendment to the existing Crit-COV-U protocol, granted by the German-Saxonian Board of Physicians (Dresden, Germany; number EK-BR-88/20.1) and the institutional review boards of the participating sites. UriCoV represents a multinational cohort study conducted in seven European countries (Austria, France, Germany, North Macedonia, Poland, Spain, Sweden), aiming at phenotyping of post-acute sequelae of SARS-CoV-2 infection (PASC) (I), comprehending PASC pathophysiology (II), and development of clinically applicable secondary prevention methods (III) (e.g., disease-specific diagnostic models via urinary peptidomics profiling (UPP)).

In 2023, based on the Delphi Consensus criteria [2], a standardized questionnaire was designed by UriCoV members, to assist in identifying participants of Crit-CoV-U [1] who had had contracted symptomatic PASC. These Crit-CoV-U participants were patients with PCR-confirmed SARS-CoV-2 infection, diagnosed either in ambulatory care or on the first day of hospital admission. Contact information was extracted from the electronic hospital registry data upon obtaining ethical clearance. The questionnaire was translated verbatim by medical professionals into six languages and disseminated to participants' residential addresses. Alternatively, the participants were contacted via phone calls and invited to participate in a telephone survey led by medical research assistants. UriCoV participants were enrolled upon positive completion of the questionnaire between January 2023 and July 2024. The collected data were entered and stored in a REDCap database [3, 4].

Our definition of PASC based on the Delphi Consensus criteria [2] was determined by responses to the first four yes/no questions in the questionnaire derived from the definition of WHO [2], which assessed: 1) whether symptoms or complaints persisted since the initial acute COVID-19 infection; 2) whether new complaints emerged within three months following the first acute COVID-19 infection; 3) whether any symptoms or complaints resulted in new health limitations; and 4) whether a pre-existing underlying condition worsened since the initial COVID-19 infection. A "yes" response to any of these questions and a symptom duration of at least two months classified patients as having had PASC. Individuals without symptoms served as controls. The urine samples were collected at a median of 1098 (IQR: 784-1533) days after the first PCR-confirmed COVID-19. All participants provided written informed consent and were non-anuric adults (≥18 years). Flowchart for the UriCoV cohort is presented in Figure E1.

#### 1.2 PASC-specific biomarker definition

##### 1.2.1 Sample preparation

The second or third urine sample of the morning was collected and stored at -20°C until shipping on dry ice. Before the analysis, urine was defrosted, and 700 µl of urine was blended with 700 µl of 2 M urea, 10 mM $NH_4OH$, and 0.02 %SDS. To eliminate high molecular weight proteins, Centristat 20 kDa cut-off centrifugal filter device ultrafiltered each solution for 1 h at 4°C. Desalination of the 1100 µl filtrate was achieved via a pre-equilibrated PD-10 gel filtration column (GE Healthcare, Uppsala, Sweden), eluted in 0.01% aqueous $NH_4OH$, resulting in concentration of the polypeptides followed by a final lyophilization process. The eluates were then stored at 4°C until the capillary electrophoresis (CE) was performed using a Programmable/Advanced Capillary Electrophoresis Multidimensional Quality (P/ACE MDQ, Beckman Coulter, Fullerton, CA) coupled on-line to MicrOTOF II mass spectrometry (MS, Bruker Daltonic, Bremen, Germany). Following the re-dispersion of samples in 10 µl high-performance liquid chromatography (HPLC)-grade water, they were applied to CE-MS with injection volumes of approximately 280 nL (2.0 psi for 99 sec). Peptide separation was conducted with reverse polarity at 25 kV. The running buffer was prepared by combining 20% $C_2H_3N$ (Sigma-Aldrich, Taufkirchen, Germany) in HPLC-grade $H_2O$ (Roth, Karlsruhe, Germany) with 0.94% $CH_2O_2$ (Sigma-Aldrich).

##### 1.2.2 Urinary peptidome analysis

With a set potential between -4.0 to -4.5 kV, spectra were acquired over an m/z range of 350-3000, accumulating every 3 sec using the electrospray ionization sprayer (ESI, Agilent Technologies). Ion peaks in the mass spectrum were then resolved into single masses using MosaFinder software [5]. For noise filtering, the analysis was restricted to signals with $z > 1$, which appeared in at least three consecutive spectra. The obtained molecular mass (Da) and capillary electrophoresis migration time (in minutes) were subsequently adjusted using local and global linear regression derived from 3151 internal standards, followed by a signal density calibration using reference signals from 29 abundant peptides. Data were transferred to a Microsoft SQL Server for further evaluation and statistical analysis.

##### 1.2.3 Sequencing of peptides

Peptide sequencing was performed using a Dionex Ultimate 3000 RSLS nanoflow system (Dionex, Camberly, UK) for LC-MS/MS analysis or a P/ACE MDQ capillary electrophoresis system (Beckman Coulter, Fullerton, CA) for CE-MS/MS analysis, both coupled to a Q Exactive™ Plus Hybrid Quadrupole-Orbitrap™ Mass Spectrometer (Thermo Scientific, Waltham, MA). The resulting spectra were searched against the UniProt human non-redundant database with Proteome Discoverer 2.4 (Thermo Fisher Scientific) software under the following parameters: high-energy collision-induced dissociation activation, 5 ppm precursor mass tolerance, 0.05 Da fragment mass tolerance, no enzyme specificity, no fixed modification, and variable modifications for oxidation (proline and methionine) and deamidation. The cross-correlation threshold was set at (Xcorr) $\geq$ 1.9. The correlation between the peptide charge at pH of 2 and CE-migration time [6], as well as the comparison of CE-migration time to the calculated migration time, served as criteria when minimizing false-positive peptide assignments.

##### 1.2.4 Statistical analysis and biomarker classifier generation

The Innsbruck cohort was used for biomarker definition, classifier generation and independent validation. Only peptides present with a frequency of at least 80% in exactly one group (case or control) were considered for further analysis. The Wilcoxon rank sum test was applied to detect potential peptide markers in the training set. Obtained p-values were corrected for multiple testing at the level of 5% according to the Benjamini–Hochberg (BH) procedure [7]. Peptides deemed statistically significant with available amino acid sequence information were included for classifier development using a support vector machine in MosaCluster software (version 1.6.5). The model was optimized through iterative selection within a leave-one-out cross-validation (LOOCV) framework on the training dataset and reduced to a final set of peptide biomarkers. A final biomarker score was assigned based on the Euclidean distance of the vector to the separating hyperplane in multidimensional space.

### 2 Supplemental Results

#### 2.1 UriCoV cohort

Of 786 questionnaires distributed to nine centers, 320 were completed, corresponding to a response rate of 41%. Among respondents, 179 reported having had or still experiencing PASC, while 139 did not at a median follow-up of 2.86 [IQR: 2.61–3.18] years. Two participants could not be classified due to missing responses. The symptom duration distributions are presented in Table E1.

#### 2.2 Application of PASC195 to the UriCoV cohort

A total of 109 participants provided urine samples, of whom 35 were identified as having persistent PASC (defined by the presence of symptoms at the time of questionnaire completion) and 40 as non-PASC. To evaluate the diagnostic performance in an external cohort with distinct clinical characteristics (population composition and sample collection time) and methodological approaches, including differences in the diagnostic criteria (self-diagnosis vs. clinician-confirmed) and questionnaire design, PASC195 was applied to UriCoV data. Sensitivity analysis yielded no statistical significance ($p = 0.1492$), with PASC195 achieving an AUC under the ROC of 0.598 (95% CI 0.465–0.731) (Figure E2).

#### 4 Supplemental Tables & Figures

**Figure 1: Flowchart for the UriCoV cohort**

*Samples were collected from centers in four countries, including Spain (n=14), Poland (n=62), France (n=18) and Sweden (n=15). Patients with symptoms persisting at the time of questionnaire completion were classified as cases (n=34), and patients who have not indicated symptoms were selected as controls (n=40).*

**Figure 2: Sensitivity analysis in the validation set of UriCoV**

*Legend: Receiver Operating Characteristic (ROC) curve visualizes the performance of PASC195 in distinguishing between PASC (n=34) and controls (n=40) within the UriCoV cohort. The bold black line represents the ROC curve. The gray dotted line represents the 95% confidence intervals.*

**Figure 3: Peptidome profile of all collagenous peptides in the PASC195 classifier**

*Legend: UniProt symbols corresponding to each peptide are displayed along the y-axis. Log2-transformed peptide signal intensities were Z-normalized and hierarchically clustered within four subgroups: PASC-ME/CFS, PASC, healthy controls, and non-COVID-19 ME/CFS. The amino acid start and end positions are indicated in parentheses. Panel A shows the peptides corresponding to collagen type 1, Panel B shows collagen type 2-4, and Panel C displays all the other collagenous peptides. Blue indicates lower signal intensities, pink indicates higher signal intensities, and gray indicates missing values. PASC: Post-acute sequelae of SARS-CoV-2 infection. ME/CFS: myalgic encephalomyelitis/chronic fatigue syndrome.*

**Table 1: Classification and urine sample collection (UriCoV)**

| Symptom duration | Questionnaires | Urine samples |
|---|---|---|
| | **Number (%)** | **Number (%)** |
| Non-PASC | 139 (43.7%) | 40 (38.5%) |
| <4 weeks | 12 (3.8%) | 8 (7.7%) |
| 4 weeks < 8 weeks | 13 (4.1%) | 4 (3.8%) |
| 8 weeks < 6 months | 18 (5.7%) | 5 (4.8%) |
| 6 months < 12 months | 12 (3.8%) | 8 (7.7%) |
| 12 months < 24 months | 9 (2.8%) | 4 (3.8%) |
| 24 months < lasting until the day of the questionnaire completion | 104 (32.7%) | 35* (33.7%) |

*Table showing stratification of patients based on self-reported symptom duration. * One sample was excluded owing to a technical error.*

**Table 2: In silico intervention result.**

| Evaluation ID | Group | PASC195 score | min_PASC195 score after intervention | predicted score change | optimal therapy |
|---|---|---|---|---|---|
| 266333 | PASC_test | 1.504 | 1.407 | -0.097 | EG |
| 266338 | PASC_test | 0.828 | 0.716 | -0.112 | EG |
| 266608 | PASC_test | 1.56 | 1.459 | -0.101 | EG |
| 266617 | PASC_test | 0.608 | 0.501 | -0.107 | EG |
| 266618 | PASC_test | -0.185 | -0.429 | -0.244 | EG |
| 267496 | PASC_test | 1.012 | 0.909 | -0.103 | EG |
| 267503 | PASC_test | 0.588 | 0.476 | -0.112 | EG |
| 266501 | PASC_test | 0.666 | 0.527 | -0.139 | EGA |
| 266597 | PASC_test | 1.133 | 1.029 | -0.104 | EGA |
| 266616 | PASC_test | 1.286 | 1.15 | -0.136 | EGA |
| 267500 | PASC_test | -0.918 | -0.989 | -0.071 | EGA |
| 266609 | PASC_test | 0.43 | 0.351 | -0.079 | G |
| 266251 | PASC_test | 0.024 | -0.122 | -0.146 | MEG |
| 266253 | PASC_test | 1.307 | 1.177 | -0.13 | MEG |
| 266267 | PASC_test | 0.721 | 0.461 | -0.26 | MEG |
| 266496 | PASC_test | 0.643 | 0.51 | -0.133 | MEG |
| 266499 | PASC_test | 1.069 | 0.948 | -0.121 | MEG |
| 266604 | PASC_test | 0.557 | 0.38 | -0.177 | MEG |
| 266607 | PASC_test | 0.854 | 0.743 | -0.111 | MEG |
| 266495 | PASC_test | 0.883 | 0.734 | -0.149 | MEGA |
| 266334 | PASC_training | 1 | 0.846 | -0.154 | EG |
| 266344 | PASC_training | 1.107 | 1.014 | -0.093 | EG |
| 266347 | PASC_training | 1.203 | 1.082 | -0.121 | EG |
| 266353 | PASC_training | 1.054 | 0.936 | -0.118 | EG |
| 266403 | PASC_training | 1 | 0.899 | -0.101 | EG |
| 266406 | PASC_training | 1 | 0.87 | -0.13 | EG |
| 266610 | PASC_training | 1 | 0.894 | -0.106 | EG |

| | | | | | |
|---|---|---|---|---|---|
| 266614 | PASC_training | 1 | 0.912 | -0.088 | EG |
| 266639 | PASC_training | 1 | 0.888 | -0.112 | EG |
| 266342 | PASC_training | 1 | 0.861 | -0.139 | EGA |
| 266434 | PASC_training | 1.069 | 0.929 | -0.14 | EGA |
| 266600 | PASC_training | 1 | 0.862 | -0.138 | EGA |
| 266603 | PASC_training | 1.053 | 0.907 | -0.146 | EGA |
| 266619 | PASC_training | 1 | 0.925 | -0.075 | EGA |
| 267497 | PASC_training | 1 | 0.865 | -0.135 | EGA |
| 267499 | PASC_training | 1.25 | 1.11 | -0.14 | EGA |
| 266335 | PASC_training | 1.407 | 1.168 | -0.239 | MEG |
| 266336 | PASC_training | 1.275 | 1.15 | -0.125 | MEG |
| 266339 | PASC_training | 1.247 | 1.137 | -0.11 | MEG |
| 266343 | PASC_training | 1.228 | 1.108 | -0.12 | MEG |
| 266345 | PASC_training | 1.431 | 1.329 | -0.102 | MEG |
| 266433 | PASC_training | 1.42 | 1.314 | -0.106 | MEG |
| 266497 | PASC_training | 1.254 | 1.138 | -0.116 | MEG |
| 266498 | PASC_training | 1.634 | 1.487 | -0.147 | MEG |
| 266500 | PASC_training | 1 | 0.774 | -0.226 | MEG |
| 266502 | PASC_training | 1 | 0.743 | -0.257 | MEG |
| 266598 | PASC_training | 1.41 | 1.302 | -0.108 | MEG |
| 266601 | PASC_training | 1.404 | 1.275 | -0.129 | MEG |
| 266612 | PASC_training | 1.013 | 0.906 | -0.107 | MEG |
| 266615 | PASC_training | 1 | 0.926 | -0.074 | MEG |

*M: mineralocorticoid receptor antagonists (MRA), G: glucagon-like peptide-1 receptor agonists (GLP-1 RA), A: angiotensin II receptor blockers (ARBs) and E: physical activity*

| Symptom | Overall N = 58 | Cases | | | Controls | | | Symptom duration in days | |
|---|---|---|---|---|---|---|---|---|---|
| | | Overall N = 47 | PASC N = 42 | PASC-ME/CFS N = 5 | Overall N = 11 | ME/CFS N = 3 | Healthy N = 8 | Median (IQR25-IQR75) | Missing (%) |
| **Fatigue** | | | | | | | | 307 (176-419) | 17.1 |
| New onset fatigue | 36 (62.1%) | 33 (70.2%) | 28 (66.7%) | 5 (100%) | 3 (27.3%) | 3 (100%) | 0 (0%) | 294 (176-419) | 21.2 |
| PEM | 32 (55.2%) | 26 (55.3%) | 21 (50%) | 5 (100%) | 6 (54.5%) | 3 (100%) | 3 (37.5%) | 256 (146-395) | 26.9 |
| Worsening with exertion | 40 (69%) | 35 (74.5%) | 30 (71.4%) | 5 (100%) | 5 (45.5%) | 3 (100%) | 2 (25%) | 298 (172-433) | 20 |
| **Sleep dysfunction** | | | | | | | | 268 (160-349) | 15.8 |
| Sleep onset difficulties | 15 (25.9%) | 13 (27.7%) | 11 (26.2%) | 2 (40%) | 2 (18.2%) | 1 (33.3%) | 1 (12.5%) | 291 (155-360) | 38.5 |
| Sleep maintenance difficulties | 32 (55.2%) | 27 (57.4%) | 24 (57.1%) | 3 (60%) | 5 (45.5%) | 2 (66.7%) | 3 (37.5%) | 268 (141-340) | 18.5 |
| Circadian rhythm disorder | 12 (20.7%) | 8 (17%) | 7 (16.7%) | 1 (20%) | 4 (36.4%) | 3 (100%) | 1 (12.5%) | 278 (216-392) | 25 |
| Unrefreshing sleep | 30 (51.7%) | 26 (55.3%) | 21 (50%) | 5 (100%) | 4 (36.4%) | 3 (100%) | 1 (12.5%) | 326 (193-424) | 23.1 |
| **Pain** | | | | | | | | 329 (258-428) | 23.1 |
| Joint pain | 27 (47.4%) | 23 (50%) | 18 (43.9%) | 5 (100%) | 4 (36.4%) | 2 (66.7%) | 2 (25%) | 344 (272-424) | 25 |
| Muscle pain | 30 (52.6%) | 25 (54.3%) | 20 (48.8%) | 5 (100%) | 5 (45.5%) | 3 (100%) | 2 (25%) | 325 (216-418) | 26.9 |
| Headache | 32 (56.1%) | 27 (58.7%) | 22 (53.7%) | 5 (100%) | 5 (45.5%) | 2 (66.7%) | 3 (37.5%) | 334 (260-424) | 28.6 |
| **Neurological/Cognitive Manifestations** | | | | | | | | 294 (172-411) | 16.3 |
| Concentration/Short-term memory impairment | 44 (75.9%) | 39 (83%) | 34 (81%) | 5 (100%) | 5 (45.5%) | 3 (100%) | 2 (25%) | 303 (186-426) | 20.5 |
| Information processing difficulties | 30 (51.7%) | 27 (57.4%) | 22 (52.4%) | 5 (100%) | 3 (27.3%) | 3 (100%) | 0 (0%) | 286 (198-423) | 22.2 |

| Symptom | Overall N = 58 | Cases | | | Controls | | | Symptom duration in days | |
|---|---|---|---|---|---|---|---|---|---|
| | | Overall N = 47 | PASC N = 42 | PASC-ME/CFS N = 5 | Overall N = 11 | ME/CFS N = 3 | Healthy N = 8 | Median (IQR25-IQR75) | Missing (%) |
| Word retrieval | 29 (50%) | 25 (53.2%) | 20 (47.6%) | 5 (100%) | 4 (36.4%) | 3 (100%) | 1 (12.5%) | 329 (260-433) | 20 |
| Reading disorder | 21 (36.2%) | 18 (38.3%) | 14 (33.3%) | 4 (80%) | 3 (27.3%) | 2 (66.7%) | 1 (12.5%) | 306 (166-376) | 33.3 |
| Sensory disturbances | 21 (36.2%) | 19 (40.4%) | 16 (38.1%) | 3 (60%) | 2 (18.2%) | 2 (66.7%) | 0 (0%) | 311 (164-395) | 21.1 |
| Disorientation | 12 (20.7%) | 11 (23.4%) | 9 (21.4%) | 2 (40%) | 1 (9.1%) | 1 (33.3%) | 0 (0%) | 429 (124-501) | 36.4 |
| Motor incoordination | 13 (22.4%) | 10 (21.3%) | 7 (16.7%) | 3 (60%) | 3 (27.3%) | 3 (100%) | 0 (0%) | 326 (254-352) | 30 |
| Overload leading to anxiety | 33 (56.9%) | 30 (63.8%) | 25 (59.5%) | 5 (100%) | 3 (27.3%) | 3 (100%) | 0 (0%) | 311 (200-437) | 23.3 |
| **Autonomic manifestations** | | | | | | | | 311 (176-415) | 18.6 |
| NHM | 28 (49.1%) | 25 (54.3%) | 20 (48.8%) | 5 (100%) | 3 (27.3%) | 1 (33.3%) | 2 (25%) | 342 (259-472) | 26.9 |
| POTS | 11 (19%) | 10 (21.3%) | 8 (19%) | 2 (40%) | 1 (9.1%) | 1 (33.3%) | 0 (0%) | 175 (124-302) | 30 |
| Light-headedness | 24 (41.4%) | 21 (44.7%) | 17 (40.5%) | 4 (80%) | 3 (27.3%) | 3 (100%) | 0 (0%) | 286 (188-395) | 28.6 |
| Extreme pallor | 9 (15.5%) | 7 (14.9%) | 6 (14.3%) | 1 (20%) | 2 (18.2%) | 2 (66.7%) | 0 (0%) | 238 (110-419) | 42.9 |
| IBS | 25 (43.1%) | 20 (42.6%) | 18 (42.9%) | 2 (40%) | 5 (45.5%) | 2 (66.7%) | 3 (37.5%) | 329 (230-412) | 30 |
| Bladder dysfunction | 10 (17.2%) | 8 (17%) | 6 (14.3%) | 2 (40%) | 2 (18.2%) | 1 (33.3%) | 1 (12.5%) | 361 (238-407) | 37.5 |
| Palpitations | 27 (46.6%) | 23 (48.9%) | 18 (42.9%) | 5 (100%) | 4 (36.4%) | 2 (66.7%) | 2 (25%) | 342 (152-437) | 34.8 |
| Exertional dyspnea | 27 (46.6%) | 24 (51.1%) | 19 (45.2%) | 5 (100%) | 3 (27.3%) | 3 (100%) | 0 (0%) | 262 (175-423) | 29.2 |
| **Neuroendocrine manifestations** | | | | | | | | 303 (176-384) | 16.7 |
| Thermostatic imbalance | 15 (25.9%) | 11 (23.4%) | 10 (23.8%) | 1 (20%) | 4 (36.4%) | 3 (100%) | 1 (12.5%) | 262 (114-429) | 18.2 |

| Symptom | Overall N = 58 | Cases | | | Controls | | | Symptom duration in days | |
|---|---|---|---|---|---|---|---|---|---|
| | | Overall N = 47 | PASC N = 42 | PASC-ME/CFS N = 5 | Overall N = 11 | ME/CFS N = 3 | Healthy N = 8 | Median (IQR25-IQR75) | Missing (%) |
| Excessive sweating | 23 (39.7%) | 20 (42.6%) | 15 (35.7%) | 5 (100%) | 3 (27.3%) | 3 (100%) | 0 (0%) | 326 (217-437) | 25 |
| Intolerance of heat/cold | 15 (25.9%) | 11 (23.4%) | 8 (19%) | 3 (60%) | 4 (36.4%) | 3 (100%) | 1 (12.5%) | 342 (175-429) | 18.2 |
| Cold extremities | 24 (41.4%) | 22 (46.8%) | 17 (40.5%) | 5 (100%) | 2 (18.2%) | 2 (66.7%) | 0 (0%) | 311 (238-361) | 22.7 |
| Weight gain | 9 (15.5%) | 7 (14.9%) | 5 (11.9%) | 2 (40%) | 2 (18.2%) | 2 (66.7%) | 0 (0%) | 352 (334-424) | 14.3 |
| Weight loss | 16 (27.6%) | 14 (29.8%) | 13 (31%) | 1 (20%) | 2 (18.2%) | 2 (66.7%) | 0 (0%) | 311 (153-429) | 35.7 |
| Stress intolerance | 41 (71.9%) | 36 (78.3%) | 31 (75.6%) | 5 (100%) | 5 (45.5%) | 3 (100%) | 2 (25%) | 306 (183-419) | 18.9 |
| **Immune manifestations** | | | | | | | | 325 (238-429) | 19.4 |
| Tender lymph nodes | 8 (13.8%) | 6 (12.8%) | 5 (11.9%) | 1 (20%) | 2 (18.2%) | 1 (33.3%) | 1 (12.5%) | 325 (220-334) | 50 |
| Sore throat | 15 (25.9%) | 13 (27.7%) | 10 (23.8%) | 3 (60%) | 2 (18.2%) | 2 (66.7%) | 0 (0%) | 344 (316-412) | 23.1 |
| Allergies | 9 (15.5%) | 6 (12.8%) | 5 (11.9%) | 1 (20%) | 3 (27.3%) | 2 (66.7%) | 1 (12.5%) | 247 (228-331) | 33.3 |
| Flu-like symptoms | 20 (34.5%) | 16 (34%) | 12 (28.6%) | 4 (80%) | 4 (36.4%) | 3 (100%) | 1 (12.5%) | 306 (177-378) | 25 |
| Increased sensitivity | 14 (24.1%) | 9 (19.1%) | 8 (19%) | 1 (20%) | 5 (45.5%) | 3 (100%) | 2 (25%) | 256 (220-281) | 22.2 |

**Table 3: Response distribution and symptom duration among questionnaire respondents**

*ME/CFS: myalgic encephalomyelitis/chronic fatigue syndrome. PASC: Post-acute sequelae of SARS-CoV-2 infection. PEM: Post-Exertional Malaise. NHM: Delayed postural hypertension or neurally mediated hypotension. POTS: Postural orthostatic tachycardia syndrome. IBS: Irritable bowel syndrome.*

261 ## Table 4: Statistical Comparison of Subgroups

| Symptom | PASC-ME/CFS vs Healthy | | | ME/CFS vs Healthy | | | PASC vs Healthy | | |
|---|---|---|---|---|---|---|---|---|---|
| | p | p_adj | Φ | p | p_adj | φ | p | p_adj | φ |
| New onset fatigue | 0.000777 | 1 | 0.004921 | 0.00606061 | 1 | 0.03290043 | 0.00059561 | 0.492 | 0.02263316 |
| PEM | 0.07536908 | 0.625 | 0.12452282 | 0.18181818 | 0.559 | 0.25589226 | 0.70410688 | 0.092 | 0.83612692 |
| Worsening with exertion | 0.02097902 | 0.732 | 0.04982517 | 0.06060606 | 0.671 | 0.0959596 | 0.01912885 | 0.355 | 0.10384232 |
| Sleep onset difficulties | 0.51048951 | 0.318 | 0.62576134 | 0.49090909 | 0.241 | 0.5486631 | 0.66062173 | 0.118 | 0.8160976 |
| Sleep maintenance difficulties | 0.59207459 | 0.22 | 0.70308858 | 0.54545455 | 0.261 | 0.56019656 | 0.44423553 | 0.144 | 0.73385995 |
| Circadian rhythm disorder | 1 | 0.101 | 1 | 0.02424242 | 0.81 | 0.07086247 | 1 | 0.042 | 1 |
| Unrefreshing sleep | 0.004662 | 0.854 | 0.02214452 | 0.02424242 | 0.81 | 0.07086247 | 0.06379838 | 0.277 | 0.2203944 |
| Joint pain | 0.02097902 | 0.732 | 0.04982517 | 0.49090909 | 0.386 | 0.5486631 | 0.44464196 | 0.142 | 0.73385995 |
| Muscle pain | 0.02097902 | 0.732 | 0.04982517 | 0.06060606 | 0.671 | 0.0959596 | 0.26885712 | 0.177 | 0.60097475 |
| Headache | 0.07536908 | 0.625 | 0.12452282 | 0.54545455 | 0.261 | 0.56019656 | 0.46349049 | 0.119 | 0.73385995 |
| Concentration/Short-term memory impairment | 0.02097902 | 0.732 | 0.04982517 | 0.06060606 | 0.671 | 0.0959596 | 0.00375959 | 0.457 | 0.05103556 |
| Information processing difficulties | 0.000777 | 1 | 0.004921 | 0.00606061 | 1 | 0.03290043 | 0.00638482 | 0.387 | 0.06065582 |
| Word retrieval | 0.004662 | 0.854 | 0.02214452 | 0.02424242 | 0.81 | 0.07086247 | 0.1167402 | 0.261 | 0.31686625 |
| Reading disorder | 0.03185703 | 0.675 | 0.06327006 | 0.15151515 | 0.542 | 0.22144522 | 0.40742781 | 0.167 | 0.73385995 |
| Sensory disturbances | 0.03496503 | 0.693 | 0.06327006 | 0.05454545 | 0.77 | 0.0959596 | 0.04293432 | 0.299 | 0.16315043 |
| Disorientation | 0.12820513 | 0.539 | 0.18737673 | 0.27272727 | 0.516 | 0.35736677 | 0.32162058 | 0.204 | 0.64882453 |
| Motor incoordination | 0.03496503 | 0.693 | 0.06327006 | 0.00606061 | 1 | 0.03290043 | 0.57985143 | 0.176 | 0.8160872 |
| Overload leading to anxiety | 0.000777 | 1 | 0.004921 | 0.00606061 | 1 | 0.03290043 | 0.00402912 | 0.436 | 0.05103556 |

| Symptom | PASC-ME/CFS vs Healthy | | | ME/CFS vs Healthy | | | PASC vs Healthy | | |
|---|---|---|---|---|---|---|---|---|---|
| | **p** | **p_adj** | **Φ** | **p** | **p_adj** | **φ** | **p** | **p_adj** | **φ** |
| NHM | 0.02097902 | 0.732 | 0.04982517 | 1 | 0.083 | 1 | 0.26885712 | 0.177 | 0.60097475 |
| POTS | 0.12820513 | 0.539 | 0.18737673 | 0.27272727 | 0.516 | 0.35736677 | 0.32441226 | 0.19 | 0.64882453 |
| Light-headedness | 0.00699301 | 0.843 | 0.02952603 | 0.00606061 | 1 | 0.03290043 | 0.03927289 | 0.313 | 0.16315043 |
| Extreme pallor | 0.38461538 | 0.365 | 0.50397878 | 0.05454545 | 0.77 | 0.0959596 | 0.57174045 | 0.161 | 0.8160872 |
| IBS | 1 | 0.025 | 1 | 0.54545455 | 0.261 | 0.56019656 | 1 | 0.04 | 1 |
| Bladder dysfunction | 0.51048951 | 0.318 | 0.62576134 | 0.49090909 | 0.241 | 0.5486631 | 1 | 0.019 | 1 |
| Palpitations | 0.02097902 | 0.732 | 0.04982517 | 0.49090909 | 0.386 | 0.5486631 | 0.45009163 | 0.134 | 0.73385995 |
| Exertional dyspnea | 0.000777 | 1 | 0.004921 | 0.00606061 | 1 | 0.03290043 | 0.01774393 | 0.342 | 0.10384232 |
| Thermostatic imbalance | 1 | 0.101 | 1 | 0.02424242 | 0.81 | 0.07086247 | 0.66576383 | 0.1 | 0.8160976 |
| Excessive sweating | 0.000777 | 1 | 0.004921 | 0.00606061 | 1 | 0.03290043 | 0.08642566 | 0.286 | 0.27122509 |
| Intolerance of heat/cold | 0.21678322 | 0.501 | 0.30510231 | 0.02424242 | 0.81 | 0.07086247 | 1 | 0.062 | 1 |
| Cold extremities | 0.000777 | 1 | 0.004921 | 0.05454545 | 0.77 | 0.0959596 | 0.03927289 | 0.313 | 0.16315043 |
| Weight gain | 0.12820513 | 0.539 | 0.18737673 | 0.05454545 | 0.77 | 0.0959596 | 0.57737545 | 0.145 | 0.8160872 |
| Weight loss | 0.38461538 | 0.365 | 0.50397878 | 0.05454545 | 0.77 | 0.0959596 | 0.09278753 | 0.259 | 0.27122509 |
| Stress intolerance | 0.02097902 | 0.732 | 0.04982517 | 0.06060606 | 0.671 | 0.0959596 | 0.01024133 | 0.399 | 0.07783409 |
| Tender lymph nodes | 1 | 0.101 | 1 | 0.49090909 | 0.241 | 0.5486631 | 1 | -0.007 | 1 |
| Sore throat | 0.03496503 | 0.693 | 0.06327006 | 0.05454545 | 0.77 | 0.0959596 | 0.18394285 | 0.218 | 0.46598856 |
| Allergies | 1 | 0.101 | 1 | 0.15151515 | 0.542 | 0.22144522 | 1 | -0.007 | 1 |
| Flu-like symptoms | 0.03185703 | 0.675 | 0.06327006 | 0.02424242 | 0.81 | 0.07086247 | 0.66224555 | 0.134 | 0.8160976 |
| Increased sensitivity | 1 | -0.058 | 1 | 0.06060606 | 0.671 | 0.0959596 | 0.65274164 | -0.055 | 0.8160976 |

*P-values result from statistical tests comparing each subgroup to healthy individuals based on symptom counts assessed by Fisher's Exact Test (p_adj are corrected values according to Benjamini-Hochberg) φ: Phi coefficient.*

#### Table 5: Associations with Peptidomics-Derived Classification Scores

| Symptom – PASC195 classification scores | Overall | | | Cases | | | Controls | | |
|---|---|---|---|---|---|---|---|---|---|
| | p | p_adj | r* | p | p_adj | r* | p-value | p | r* |
| New onset fatigue | 0.060789 | 0.700134 | 0.247255 | 0.103431 | 0.561484 | 0.239229 | 0.018634 | 0.093622 | 0.740233 |
| PEM | 0.87575 | 0.953239 | 0.021557 | 0.403959 | 0.769759 | 0.123295 | 0.054681 | 0.143903 | 0.606911 |
| Worsening with exertion | 0.59649 | 0.903696 | 0.070624 | 0.549991 | 0.820939 | 0.088974 | 0.022174 | 0.093622 | 0.717258 |
| Sleep onset difficulties | 0.650664 | 0.903696 | 0.060627 | 0.490361 | 0.820939 | 0.102343 | 0.905973 | 0.930458 | 0.071229 |
| Sleep maintenance difficulties | 0.78437 | 0.953239 | 0.036956 | 0.377652 | 0.769759 | 0.130256 | 0.714379 | 0.754067 | 0.137934 |
| Circadian rhythm disorder | 0.744171 | 0.953239 | 0.04411 | 0.234456 | 0.669686 | 0.175492 | 0.071951 | 0.143903 | 0.571102 |
| Unrefreshing sleep | 0.646193 | 0.903696 | 0.061299 | 0.513964 | 0.820939 | 0.096763 | 0.029387 | 0.111672 | 0.685323 |
| Joint pain | 0.179414 | 0.845911 | 0.178885 | 0.194885 | 0.622711 | 0.192743 | 0.058207 | 0.143903 | 0.599657 |
| Muscle pain | 0.148051 | 0.845911 | 0.192646 | 0.063948 | 0.561484 | 0.274768 | 0.16993 | 0.222667 | 0.44139 |
| Headache | 0.961519 | 0.961519 | 0.007456 | 0.946636 | 0.999435 | 0.011513 | 0.464192 | 0.518802 | 0.248282 |
| Concentration/Short-term memory impairment | 0.04369 | 0.700134 | 0.266045 | 0.031441 | 0.4524 | 0.315886 | 0.022174 | 0.093622 | 0.717258 |
| Information processing difficulties | 0.166067 | 0.845911 | 0.182875 | 0.189322 | 0.622711 | 0.19303 | 0.018634 | 0.093622 | 0.740233 |
| Word retrieval | 0.073698 | 0.700134 | 0.23586 | 0.035716 | 0.4524 | 0.307893 | 0.107395 | 0.163241 | 0.513992 |
| Reading disorder | 0.364898 | 0.845911 | 0.120035 | 0.26435 | 0.669686 | 0.164406 | 0.21962 | 0.260799 | 0.40096 |
| Sensory disturbances | 0.87784 | 0.953239 | 0.021245 | 0.922285 | 0.999435 | 0.015811 | 0.098193 | 0.155472 | 0.534217 |

| Symptom – PASC195 classification scores | Overall | | | Cases | | | Controls | | |
|---|---|---|---|---|---|---|---|---|---|
| | p | p_adj | r* | p | p_adj | r* | p-value | p | r* |
| Disorientation | 0.265546 | 0.845911 | 0.147454 | 0.333342 | 0.769759 | 0.142943 | 0.204871 | 0.251132 | 0.430037 |
| Motor incoordination | 0.229083 | 0.845911 | 0.15915 | 0.755084 | 0.956439 | 0.047397 | 0.018634 | 0.093622 | 0.740233 |
| Overload leading to anxiety | 0.326405 | 0.845911 | 0.129893 | 0.425393 | 0.769759 | 0.117884 | 0.018634 | 0.093622 | 0.740233 |
| NHM | 0.334158 | 0.845911 | 0.128977 | 0.588973 | 0.828925 | 0.081292 | 0.682408 | 0.7409 | 0.154215 |
| POTS | 0.13948 | 0.845911 | 0.195338 | 0.164302 | 0.622711 | 0.204757 | 0.204871 | 0.251132 | 0.430037 |
| Light-headedness | 0.487248 | 0.894203 | 0.092254 | 0.368776 | 0.769759 | 0.13266 | 0.018634 | 0.093622 | 0.740233 |
| Extreme pallor | 0.408323 | 0.862015 | 0.109981 | 0.091343 | 0.561484 | 0.248452 | 0.098193 | 0.155472 | 0.534217 |
| IBS | 0.665881 | 0.903696 | 0.05773 | 0.65913 | 0.894533 | 0.065913 | 1 | 1 | 0.027587 |
| Bladder dysfunction | 0.313121 | 0.845911 | 0.1338 | 0.561695 | 0.820939 | 0.086714 | 0.156355 | 0.212196 | 0.462988 |
| Palpitations | 0.378434 | 0.845911 | 0.116676 | 0.424813 | 0.769759 | 0.117967 | 0.448659 | 0.516637 | 0.256996 |
| Exertional dyspnea | 0.047724 | 0.700134 | 0.260987 | 0.032443 | 0.4524 | 0.313544 | 0.018634 | 0.093622 | 0.740233 |
| Thermostatic imbalance | 0.563839 | 0.903696 | 0.076949 | 0.074416 | 0.561484 | 0.262062 | 0.071951 | 0.143903 | 0.571102 |
| Excessive sweating | 0.450214 | 0.894203 | 0.100188 | 0.254101 | 0.669686 | 0.16792 | 0.018634 | 0.093622 | 0.740233 |
| Intolerance of heat/cold | 0.494165 | 0.894203 | 0.09094 | 0.821087 | 0.974172 | 0.034819 | 0.071951 | 0.143903 | 0.571102 |
| Cold extremities | 0.782319 | 0.953239 | 0.037316 | 0.948999 | 0.999435 | 0.010885 | 0.098193 | 0.155472 | 0.534217 |
| Weight gain | 0.659741 | 0.903696 | 0.05922 | 0.845991 | 0.974172 | 0.030512 | 0.098193 | 0.155472 | 0.534217 |
| Weight loss | 0.903068 | 0.953239 | 0.017134 | 1 | 1 | 0.001697 | 0.044639 | 0.143903 | 0.641061 |
| Stress intolerance | 0.901061 | 0.953239 | 0.017643 | 0.831278 | 0.974172 | 0.033378 | 0.054681 | 0.143903 | 0.606911 |
| Tender lymph nodes | 0.946063 | 0.961519 | 0.010364 | 0.55533 | 0.820939 | 0.088355 | 0.156355 | 0.212196 | 0.462988 |

| **Symptom – PASC195 classification scores** | **Overall** | | | **Cases** | | | **Controls** | | |
|---|---|---|---|---|---|---|---|---|---|
| | **p** | **p_adj** | **r*** | **p** | **p_adj** | **r*** | **p-value** | **p** | **r*** |
| Sore throat | 0.207363 | 0.845911 | 0.166724 | 0.175199 | 0.622711 | 0.199482 | 0.098193 | 0.155472 | 0.534217 |
| Allergies | 0.838332 | 0.953239 | 0.0282 | 0.196646 | 0.622711 | 0.190661 | 0.065577 | 0.143903 | 0.586018 |
| Flu-like symptoms | 0.550419 | 0.903696 | 0.079481 | 0.973134 | 0.999435 | 0.00655 | 0.046719 | 0.143903 | 0.628213 |
| Increased sensitivity | 0.313202 | 0.845911 | 0.133619 | 0.715097 | 0.937024 | 0.055216 | 0.119846 | 0.17516 | 0.496564 |

*P-values result from statistical tests (Wilcoxon rank sum test) assessing the relationship between PASC195 classification scores and each symptom before correction for multiple testing (p) and after correction (p_adj, Benjamini-Hochberg). *Effect sizes were quantified using Rank-Biserial Correlation (r).*

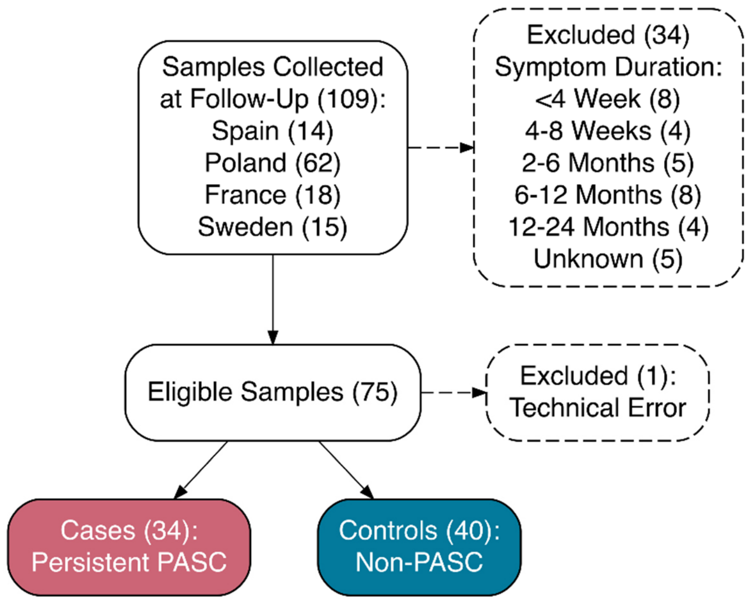


**Figure E1: Flowchart for the UriCoV cohort**

Samples were collected from centers in four countries, including Spain (n=14), Poland (n=62), France (n=18) and Sweden (n=15). Patients with symptoms persisting at the time of questionnaire completion were classified as cases (n=34), and patients who have not indicated symptoms were selected as controls (n=40).

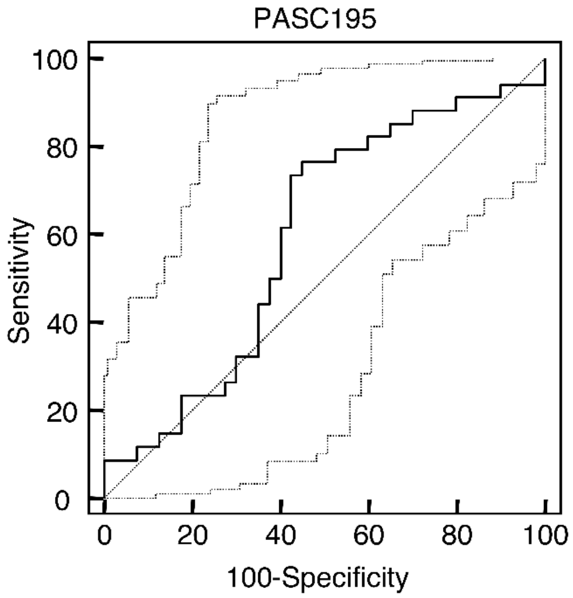


**Figure E2: Sensitivity analysis in the validation set of UriCoV**

Legend: Receiver Operating Characteristic (ROC) curve visualizes the performance of PASC195 in distinguishing between PASC (n=34) and controls (n=40) within the UriCoV cohort. The bold black line represents the ROC curve. The gray dotted line represents the 95% confidence intervals.

**A**

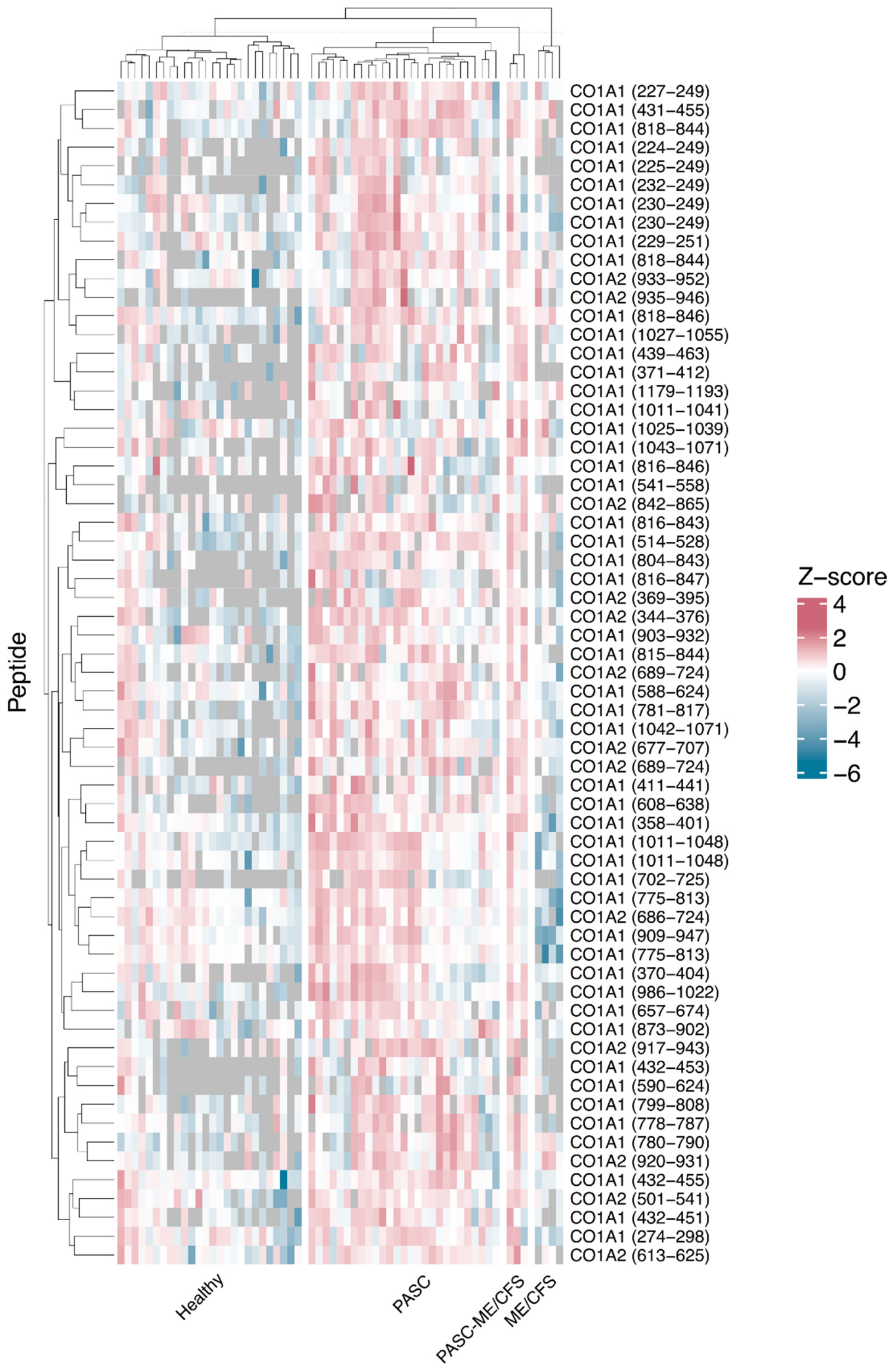
Peptide
CO1A1 (227–249)
CO1A1 (431–455)
CO1A1 (818–844)
CO1A1 (224–249)
CO1A1 (225–249)
CO1A1 (232–249)
CO1A1 (230–249)
CO1A1 (230–249)
CO1A1 (229–251)
CO1A1 (818–844)
CO1A2 (933–952)
CO1A2 (935–946)
CO1A1 (818–846)
CO1A1 (1027–1055)
CO1A1 (439–463)
CO1A1 (371–412)
CO1A1 (1179–1193)
CO1A1 (1011–1041)
CO1A1 (1025–1039)
CO1A1 (1043–1071)
CO1A1 (816–846)
CO1A1 (541–558)
CO1A2 (842–865)
CO1A1 (816–843)
CO1A1 (514–528)
CO1A1 (804–843)
CO1A1 (816–847)
CO1A2 (369–395)
CO1A2 (344–376)
CO1A1 (903–932)
CO1A1 (815–844)
CO1A2 (689–724)
CO1A1 (588–624)
CO1A1 (781–817)
CO1A1 (1042–1071)
CO1A2 (677–707)
CO1A2 (689–724)
CO1A1 (411–441)
CO1A1 (608–638)
CO1A1 (358–401)
CO1A1 (1011–1048)
CO1A1 (1011–1048)
CO1A1 (702–725)
CO1A1 (775–813)
CO1A2 (686–724)
CO1A1 (909–947)
CO1A1 (775–813)
CO1A1 (370–404)
CO1A1 (986–1022)
CO1A1 (657–674)
CO1A1 (873–902)
CO1A2 (917–943)
CO1A1 (432–453)
CO1A1 (590–624)
CO1A1 (799–808)
CO1A1 (778–787)
CO1A1 (780–790)
CO1A2 (920–931)
CO1A1 (432–455)
CO1A2 (501–541)
CO1A1 (432–451)
CO1A1 (274–298)
CO1A2 (613–625)
Z–score
4
2
0
−2
−4
−6
Healthy
PASC
PASC-ME/CFS
ME/CFS

**B**

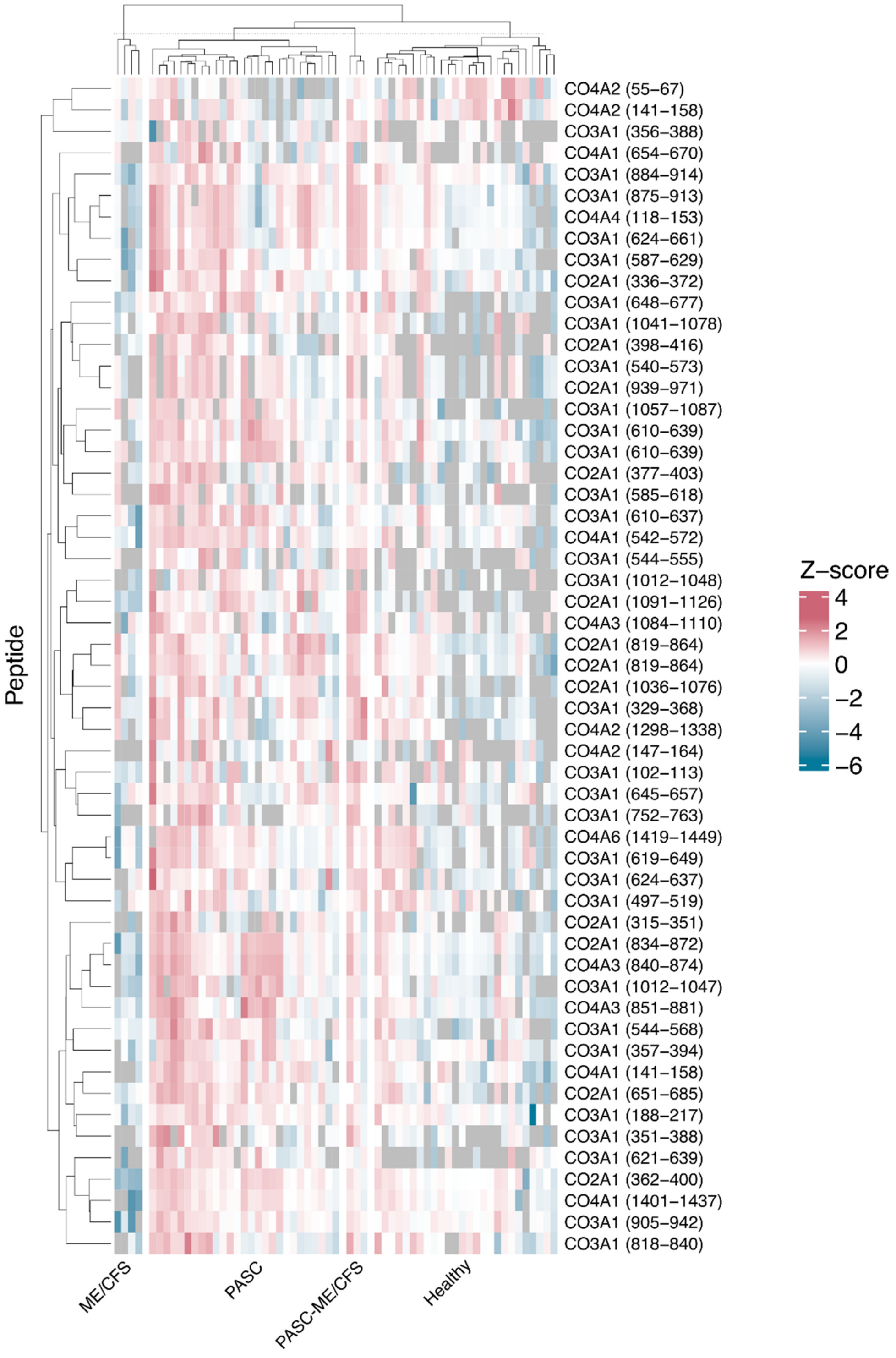
Peptide
CO4A2 (55–67)
CO4A2 (141–158)
CO3A1 (356–388)
CO4A1 (654–670)
CO3A1 (884–914)
CO3A1 (875–913)
CO4A4 (118–153)
CO3A1 (624–661)
CO3A1 (587–629)
CO2A1 (336–372)
CO3A1 (648–677)
CO3A1 (1041–1078)
CO2A1 (398–416)
CO3A1 (540–573)
CO2A1 (939–971)
CO3A1 (1057–1087)
CO3A1 (610–639)
CO3A1 (610–639)
CO2A1 (377–403)
CO3A1 (585–618)
CO3A1 (610–637)
CO4A1 (542–572)
CO3A1 (544–555)
CO3A1 (1012–1048)
CO2A1 (1091–1126)
CO4A3 (1084–1110)
CO2A1 (819–864)
CO2A1 (819–864)
CO2A1 (1036–1076)
CO3A1 (329–368)
CO4A2 (1298–1338)
CO4A2 (147–164)
CO3A1 (102–113)
CO3A1 (645–657)
CO3A1 (752–763)
CO4A6 (1419–1449)
CO3A1 (619–649)
CO3A1 (624–637)
CO3A1 (497–519)
CO2A1 (315–351)
CO2A1 (834–872)
CO4A3 (840–874)
CO3A1 (1012–1047)
CO4A3 (851–881)
CO3A1 (544–568)
CO3A1 (357–394)
CO4A1 (141–158)
CO2A1 (651–685)
CO3A1 (188–217)
CO3A1 (351–388)
CO3A1 (621–639)
CO2A1 (362–400)
CO4A1 (1401–1437)
CO3A1 (905–942)
CO3A1 (818–840)
Z–score
4
2
0
−2
−4
−6
ME/CFS
PASC
PASC-ME/CFS
Healthy

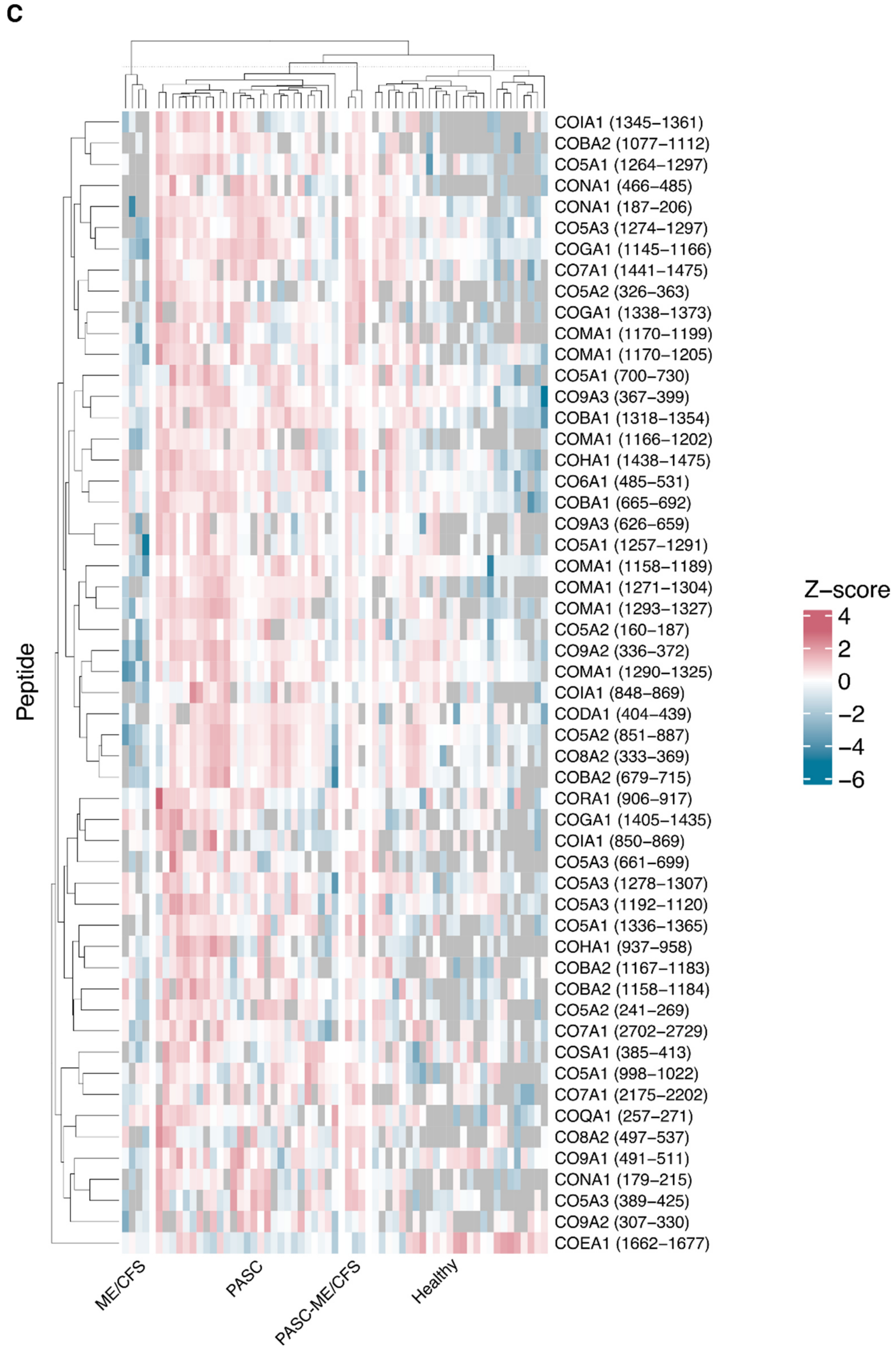


**Figure E3: Peptidome profile of all collagenous peptides in the PASC195 classifier**

Legend: UniProt symbols corresponding to each peptide are displayed along the y-axis. Log2-transformed peptide signal intensities were Z-normalized and hierarchically clustered within four subgroups: PASC-ME/CFS, PASC, healthy controls, and non-COVID-19 ME/CFS. The amino acid start and end positions are indicated in parentheses. Panel A shows the peptides corresponding to collagen type 1, Panel B shows collagen type 2-4, and Panel C displays all the other collagenous peptides. Blue indicates lower signal intensities, pink indicates higher signal intensities, and gray indicates missing values. PASC: Post-acute sequelae of SARS-CoV-2 infection. ME/CFS: myalgic encephalomyelitis/chronic fatigue syndrome.
